# HyTrax: Deep Sequential Modeling of Serial Musculoskeletal Measurements for Fracture Prediction in the Women’s Health Initiative with External Evaluation in the Framingham Heart Study

**DOI:** 10.64898/2026.06.30.26356875

**Authors:** Jongyun Jung, Qing Wu

**Affiliations:** Department of Biomedical Informatics, College of Medicine, The Ohio State University Wexner Medical Center, Columbus, Ohio, USA (Dr. Qing Wu, Jongyun Jung)

**Keywords:** Fractures, Transformer, Longitudinal trajectories, BMD, Muscle, FRAX, Aging

## Abstract

The clinical utility of monitoring longitudinal changes in musculoskeletal trajectories, including bone mineral density (BMD), muscle strength, height, and weight for fracture prediction, remains underutilized, as current gold-standard tools such as the Fracture Risk Assessment Tool (FRAX) rely solely on cross-sectional baseline data. This study aimed to determine whether a deep learning model integrating individualized musculoskeletal trajectories improves fracture prediction accuracy compared to established static benchmarks. We developed the Hybrid Trajectory-Based model (HyTrax), a Transformer-based deep learning model that encodes sequential measurements of hip and spine BMD, grip strength, height, and weight as temporal tokens, incorporating subject-specific slopes derived from linear mixed-effects models. The model was trained and internally validated in 27,512 postmenopausal women from the Women’s Health Initiative (WHI) and externally evaluated in 1,193 participants from the Framingham Heart Study (FHS). In the WHI validation set, the HyTrax + FRAX (BMD) ensemble model achieved a time-dependent Area Under the Curve (AUC) of 0.85 for Major Osteoporotic Fracture, outperforming both the longitudinal Transformer alone (AUC = 0.80) and the standard FRAX-BMD model (AUC = 0.82). The HyTrax + FRAX (BMD) ensemble model demonstrated favorable discrimination and improved risk stratification (Net Reclassification Improvement +26.5%) in WHI. Evaluation in the FHS cohort demonstrated the transportability of the longitudinal embeddings, with the HyTrax + Baseline 2 ensemble model (integrating longitudinal embeddings with clinical risk factors, BMD, and grip strength) achieving an AUC of 0.74. Explainability analyses identified early longitudinal weight fluctuations and overall height loss trajectories as important predictors of future fracture risk, alongside static factors such as age and genetic predisposition. By leveraging individualized trajectories through deep sequential modeling with baseline FRAX probability, the HyTrax + FRAX (BMD) ensemble model improved fracture discrimination over static assessments, offering a framework for incorporating repeated clinical measures into fracture prediction.

**Lay Summary:** Current fractures screening tools such as FRAX provide a risk score based on a single "snapshot" of a patient’s health, often overlooking warning signs hidden in how bodies change over time. To address this, we developed "HyTrax," an artificial intelligence model that analyzes a patient’s long-term history of bone density, muscle strength, height, and weight changes to predict future fractures. Evaluated on over 27,000 individuals, the HyTrax + FRAX (BMD) ensemble model outperformed standard tools. This trajectory-based approach offers a more accurate, personalized method for identifying high-risk patients.

## Introduction

Osteoporotic fractures are a major cause of morbidity, disability, and mortality among aging adults ^1^, and low bone mineral density (BMD) remains one of the strongest established predictors of fracture risk ^2^. However, despite the biological plausibility that greater bone loss over time elevates fracture risk, the clinical utility of monitoring longitudinal changes in BMD remains controversial ^3,4^. This uncertainty is reflected in current guidelines; the U.S. Preventive Services Task Force acknowledges limited evidence for serial BMD testing intervals, and population-based studies have yielded conflicting results regarding whether repeated measurements improve prediction accuracy beyond baseline values alone ^5,6^. Consequently, there is no consensus on how to incorporate temporal bone density changes into routine fracture risk assessment.

Concurrently, growing evidence indicates that fracture risk is driven not only by bone loss but also by parallel declines in the bone–muscle unit ^4,7^. Longitudinal deterioration in grip strength, weight loss, and height loss have all emerged as significant, independent predictors of fracture risk ^8–12^. Yet, current prediction models have failed to jointly evaluate these repeated musculoskeletal and anthropometric trajectories. The most widely used fracture risk prediction tool, the Fracture Risk Assessment Tool (FRAX) ^13^, incorporates baseline age, clinical risk factors, and optionally a single BMD value, but it does not account for longitudinal changes in body composition, bone density, or muscle strength. Furthermore, FRAX assumes uniform risk relationships across racial groups, potentially limiting accuracy in diverse populations ^14,15^. As a result, decisions about intensifying treatment often rely on subjective clinical judgment of "meaningful change" rather than data-driven longitudinal metrics ^16^.

In parallel, advances in machine learning (ML) have demonstrated potential for improving fracture prediction ^17–21^. However, prior efforts have largely relied on static baseline predictors or diagnostic codes, neglecting quantitative physiological trajectories ^22^. To date, no fracture prediction model has leveraged modern sequential architectures, such as Transformers ^23^, to analyze longitudinal musculoskeletal data. Although deep learning excels at modeling complex temporal sequences, it often lacks the robust population-level calibration of established epidemiological tools ^24,25^. Therefore, an optimal predictive framework should leverage both the individualized dynamic patterns learned from repeated measurements and the stable, population-anchored risk estimates provided by current clinical standards.

To address these gaps, we developed the Hybrid Trajectory-Based model (HyTrax), a Transformer-based deep learning model designed to learn individualized temporal dynamics that traditional models, including FRAX, cannot capture. The HyTrax architecture encodes sequential measurements of hip and spine BMD, grip strength, height, and weight as temporal tokens, incorporating subject-specific slopes derived from linear mixed-effects models. We trained and calibrated the model using 27,512 postmenopausal women from the Women’s Health Initiative (WHI) and performed external evaluation in the Framingham Heart Study (FHS). Crucially, to maximize clinical utility and predictive power, we integrated the HyTrax with the baseline FRAX probability to form the HyTrax + FRAX (BMD) ensemble model. This study evaluated whether combining longitudinal musculoskeletal measurements with established baseline risk assessment could improve 10-year fracture prediction within the study designs used in WHI and FHS data sets.

## Data Sources

### Women’s Health Initiative

#### Methodology

Model development and internal validation were performed using the WHI, a large, racially and ethnically diverse prospective cohort of postmenopausal women aged 50–79 years ^26^. We harmonized genetic and phenotypic data across six WHI sub-studies: Genomics and Randomized Trials Network (GARNET, N=4,894), SNP Health Association Resource (SHARE, N=12,007), Population Architecture using Genomics and Epidemiology (PAGE, N=12,646), Women’s Health Initiative Memory Study (WHIMS, N=5,740), WHI Sequencing Project (WHISP, N=1,904), and Genome-Wide Association Study (GWAS) Hip Fracture (N=3,688)—into a unified longitudinal analytic dataset (**Table S1**). Following exclusions for missing race/ethnicity, duplicated participants across sub-studies, osteoporosis-related medication use at the baseline visit, and rare categories, the final analytic sample included 27,512 unique participants (**Figure S1**). WHI provided repeated measurements of BMD (hip and spine), muscle strength, anthropometrics, and adjudicated fracture outcomes, enabling the development of a longitudinal deep-learning model.

#### Framingham Heart Study

External evaluation was conducted using the original cohort of the FHS, a longitudinal community-based cohort initiated in 1948 and predominantly of White European ancestry ^27,28^. Bone densitometry was first obtained at Exam 22 (1992–1993), with additional scans at Exam 24 (1996–1997). After excluding individuals lacking BMD information, 1,193 participants constituted the external evaluation dataset (**Figure S2**).

### Fracture probability and FRAX assessment

In the WHI data set, the 10-year absolute risk of major osteoporotic fracture (MOF) and hip fracture was estimated using the US FRAX model, version 3.0. We utilized pre-calculated FRAX probabilities provided within the dbGaP dataset releases, which were computed using the standard FRAX algorithm. Two distinct FRAX probabilities were evaluated for each fracture outcome: FRAX (WHO), calculated using clinical risk factors alone, and FRAX (BMD), which additionally incorporated femoral neck bone mineral density.

### Longitudinal Measurements and Covariates

Longitudinal measures included BMD (hip and spine), grip strength, height and weight. WHI assessments were aligned by visit year; FHS data were harmonized across Exam 20, 22 and 24. Static clinical risk factors (CRFs) included smoking status, alcohol intake, parental hip fracture history, rheumatoid arthritis, glucocorticoid use, and previous fragility fracture, following standard FRAX definitions ^13^. However, parental hip fracture history was exclusively available in the WHI cohort and was therefore omitted from all FHS analyses. In FHS, grip strength was available only at Exam 24; placeholder variables were created for earlier exams to preserve temporal structure.

The FHS original cohort participants underwent bone densitometry by DXA with a Lunar DPX-L (Lunar Corp., Madison, WI, USA) during their examination 22 (1992–1993). To maximize the sample size, we used DXA scans from examination 24 (in 1996–1997) for 31 Original Cohort members who did not have DXAs at examination 22. All non-anthropometric covariates were collected through self-reported questionnaires and verified through medical records when available. These included smoking status, alcohol intake, parental hip fracture history, rheumatoid arthritis, glucocorticoid use, and previous fragility fracture (with parental hip fracture history utilized only in the WHI cohort due to its unavailability in the FHS dbGaP dataset). Smoking status was recorded as “Never,” “Past,” and “Current.” Previous falls were assessed through self-report using the questionnaire item: “During the past 12 months, how many times did you fall and land on the floor or ground?” Responses were dichotomized into a binary variable indicating the presence or absence of any falls within the past 12 months (Yes/No).

### Standardization and Handling of Missing Data

To facilitate stable convergence during Transformer training, all continuous longitudinal and static predictors, including BMD, grip strength, height, weight, and age, were standardized to a mean of zero and unit variance. Standardization parameters were derived exclusively from the WHI training split and applied to the validation and FHS external testing sets to prevent data leakage. Missing data were handled using a hybrid imputation strategy tailored to the variable type, applied consistently across both the WHI and FHS cohorts. For continuous variables, missing values were imputed using Multiple Imputation by Chained Equations (MICE) with Predictive Mean Matching (PMM) to preserve non-normal data distributions ^29^. A total of m=5 complete datasets were generated for each cohort with 50 iterations per chain to ensure convergence. To appropriately account for imputation uncertainty during deep learning evaluation, we applied a rigorous per-imputation workflow. Specifically, the calibration procedure (Platt scaling) and the estimation of the ensemble weight (α), both detailed in subsequent sections, were performed independently within the calibration split of each of the m=5 imputed datasets. We then generated predictions for each participant across all 5 datasets, utilizing their respective dataset-specific models and weights. The resulting predicted probabilities were pooled (averaged) across the imputations before computing the final discrimination and calibration metrics, with uncertainty intervals subsequently derived via bootstrapping. For categorical clinical risk factors, missing values were imputed as "No" (absence of the risk factor), adhering to standard FRAX clinical guidelines ^13^.

### Fracture Outcomes and Follow-Up

To prevent data leakage and immortal time bias, all longitudinal sequences were strictly truncated prior to the outcome evaluation window. The primary outcome was incident major osteoporotic fracture (MOF), defined as a fracture at the hip, spine, shoulder, or wrist; incident hip fracture was evaluated as a secondary outcome. All outcomes were defined as incident fracture occurring within a prespecified 10-year absolute risk horizon (H = 10) following the prediction time (t₀). **Table S2** summarizes the prediction-time and evaluation design for each cohort.

To emulate a real-world clinical scenario in which a clinician uses all available longitudinal history at the time of assessment, t₀ in WHI was defined individually for each participant as their final recorded clinical visit prior to fracture, death, or censoring. All measurements after this individualized t₀ were strictly masked, ensuring no post-t₀ data were accessible to the model. Participants who experienced an incident fracture within 10 years of t₀ were classified as events. Participants who died before fracture were treated as non-events during model fitting (cause-specific approach); no formal competing-risk modeling was performed. During evaluation, model-generated probabilities were treated as continuous risk scores and all participants, including those censored before 10 years, were retained. Time-dependent survival metrics with inverse-probability-of-censoring weighting (IPCW) were applied to adjust for right-censoring across the full follow-up period.

For external evaluation in FHS, a strict fixed-landmark design was applied to test transportability without the advantage of a rolling observation window. The landmark time t₀ was set universally at Exam 24, ensuring that all longitudinal inputs (Exams 20, 22, and 24) temporally preceded the outcome window. Fractures occurring between Exam 20 and Exam 24 were classified as prevalent events and encoded as a binary covariate ("previous fragility fracture"); only fractures occurring strictly within the 10-year window after Exam 24 were counted as outcome events. The same IPCW-based evaluation framework used in WHI was applied in FHS.

### Genotyping and Polygenic Risk Score Development

WHI and FHS participants were genotyped using the Illumina (Illumina Inc., San Diego, CA, USA) or Affymetrix 6.0 Array platforms (Affymetrix Inc., Santa Clara, CA, USA). Rigorous quality control included filtering for minor allele frequency (MAF ≥ 0.01), individual missingness (<5%), and deviation from Hardy-Weinberg equilibrium (P < 0.0001) using PLINK 1.9 ^30^. Genotype imputation was performed via the Michigan Imputation Server ^31^ using the Haplotype Reference Consortium panel ^32^ and the 1000 Genomes Phase 3 (Version 5) ^33^ to ensure genomic representation across diverse populations. Polygenic Risk Score (PRS) for fracture risk was computed employing the LDpred Bayesian algorithm ^34^, utilizing summary statistics from a large-scale UK Biobank genome-wide association study involving 103,155 genetic variants ^35^. PRS was used as a static risk factor in both the WHI and FHS data sets.

### Mixed-Effects Modeling of Longitudinal Trajectories

To quantify subject-specific temporal dynamics, we fit linear mixed-effects models (LMMs) with random intercepts and slopes for each longitudinal biomarker (hip BMD, spine BMD, grip strength, height, and weight). Each model took the form:

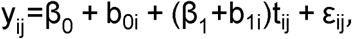

where y_ij_ and t_ij_ represent the biomarker measurement and the time for participant i at time j, respectively. The fixed effects β_0_ and β_1_ represents the population-average intercept and rate of change (slope), respectively, while b_0i_ and b_1i_ represent participant-specific deviations. The term ε_ij_ represents the residual measurement error. Valid slope estimation required a minimum of two distinct visits. For participants with sufficient longitudinal data (≥ 2 visits), subject-specific slopes (b_1i_) were derived from the LMMs Best Linear Unbiased Predictors (BLUPs). For participants with <2 visits (insufficient data for slope estimation), the subject-specific random-slope deviation b_1i_ was set to 0, meaning only the population-average slope β_1_ was applied, and a missingness indicator was included. In the WHI cohort, longitudinal hip BMD data was available for a subset of 2,669 participants; of these, 2,391 (89.6%) had ≥ 2 visits allowing for calculation of dynamic trajectories, while 278 (10.4%) had only a single baseline visit and relied on imputed slopes. Similarly, in the FHS external evaluation cohort, hip BMD data was available for 1,179 participants; of these, 786 (66.7%) had ≥ 2 visits to compute dynamic trajectories, while 393 (33.3%) had only a single visit and utilized imputed slopes (**Table S3**).

### Hybrid Trajectory-Based Model (HyTrax)

#### Tokenization of Longitudinal Inputs

At each visit, multivariate measures were encoded as baseline tokens and visit-to-visit change tokens Δ spanning from baseline (V0) through Visit 10 (V10) (**Figure 1A**). These structured tokens capture short- and long-term physiological changes, enabling the Transformer to learn temporal dependencies across visits (**Figure 1B**).

**Figure 1.**
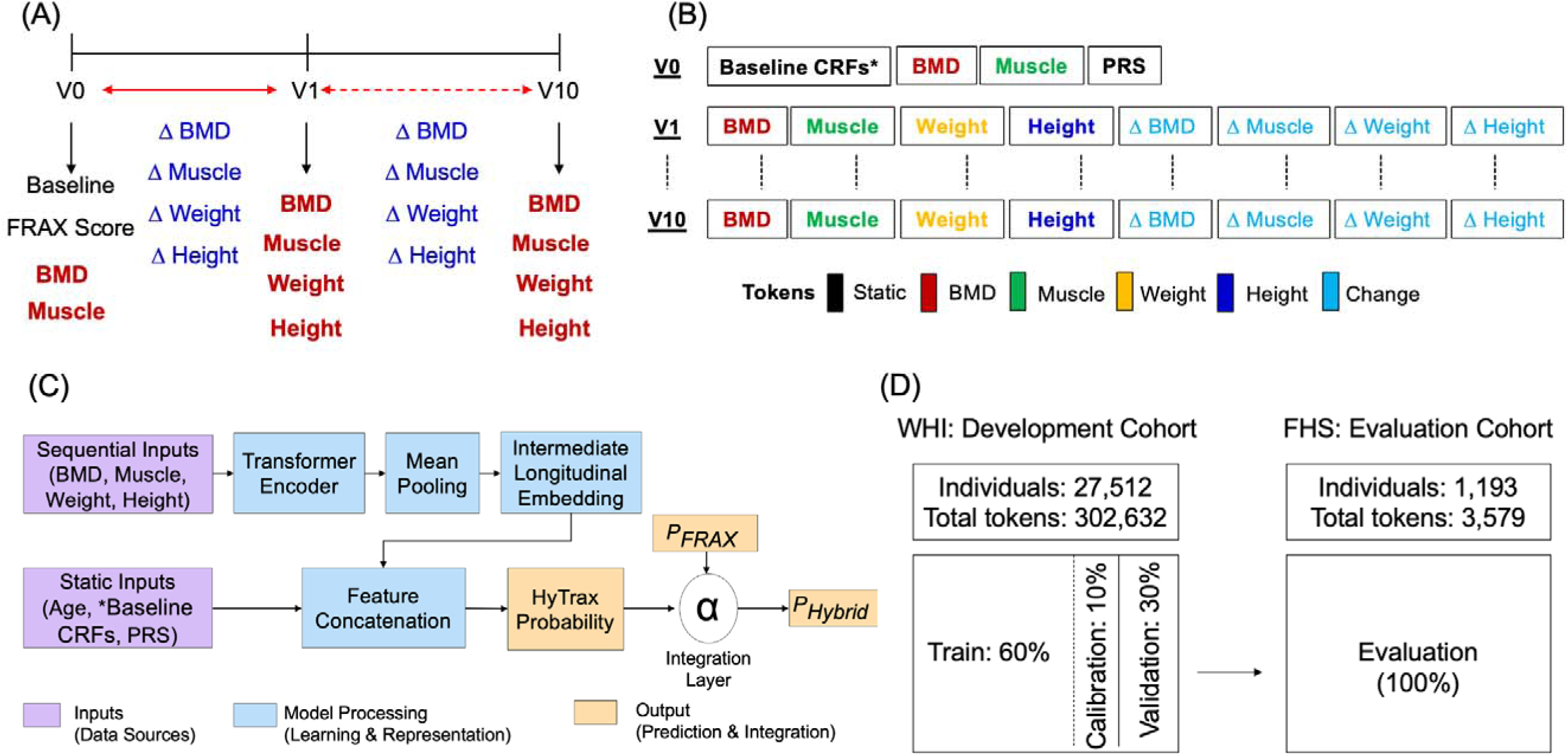
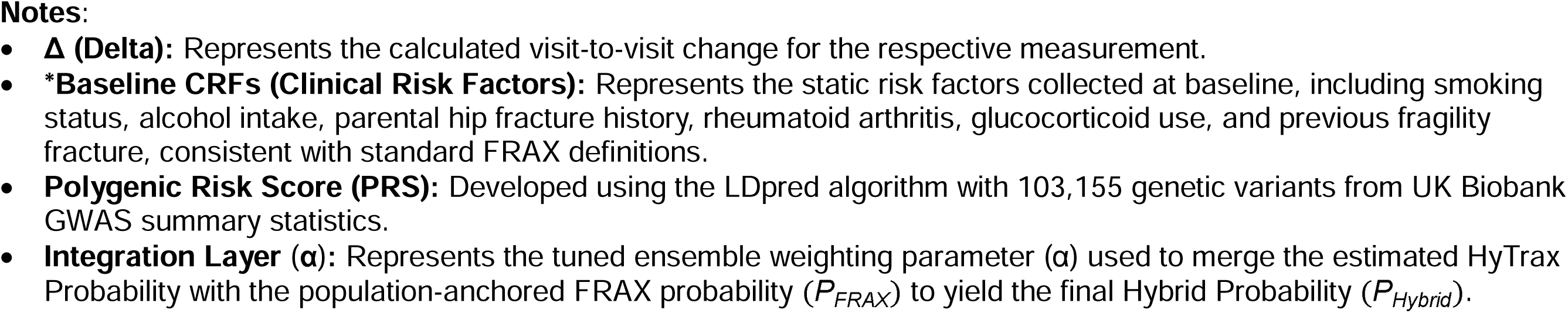
Overview of Hybrid Trajectory-Based (HyTrax) Model Inputs, Cohorts, and Modeling Framework. (A) Longitudinal inputs for bone mineral density (BMD) at hip and spine, muscle strength, weight, and height are represented using baseline values and visit-to-visit changes ( ; from baseline, V0 through Visit 10, V10) to capture temporal dynamics. (B) Longitudinal measurements are converted into tokens, where each token represents either a baseline value or a visit-to-visit change, enabling the Transformer to learn temporal dynamics across visits. (C) The HyTrax architecture integrates Transformer-derived longitudinal embeddings with static clinical covariates to generate a dynamic risk embedding, which is subsequently combined with FRAX-based risk to form the hybrid ensemble. (D) The Women’s Health Initiative (WHI, N=27,512) provides training, calibration, and internal validation data, while the Framingham Heart Study (FHS, N=1,193) enables external evaluation.

#### Model Architecture

The HyTrax uses a Transformer encoder adapted for biomedical time series (**Figure 1C**). Each visit’s feature vector is projected into a latent space and augmented with sinusoidal positional encodings ^23^. Multi-head self-attention layers capture dependencies across time, and attention-pooled temporal embeddings are concatenated with static clinical risk factors, demographic variables, and mixed-effects slopes. The combined representation is passed through a feed-forward prediction head to generate calibrated fracture risk logits.

#### Hybrid Integration with FRAX

To quantify the synergistic contribution of deep learning and established risk tools, the HyTrax was integrated with the standard clinical risk probability (FRAX) to form the HyTrax + FRAX ensemble model:

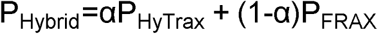

The weight parameter α was optimized independently on the calibration subset of each imputed dataset to maximize the area under the receiver operating characteristic curve (AUC). Values of α near 1 indicate dominance of the HyTrax, while values near 0 reflect reliance on FRAX. This formulation allows the HyTrax + FRAX ensemble model to leverage both the individualized temporal dynamics captured by the HyTrax and the robust population-level calibration of FRAX.

### Comparison of Models and Benchmarking

To contextualize the predictive performance of the HyTrax framework against established standards, we compared it to baseline FRAX scores, utilizing specific adaptation strategies to account for cohort differences. In the WHI internal validation, we evaluated the model against two FRAX models calculated at the baseline visit: the standard clinical risk factor model FRAX (WHO) and the model incorporating clinical risk factors and BMD (FRAX BMD). Conversely, for the external FHS evaluation where specific variables required for the official FRAX algorithm were unavailable, we developed two surrogate logistic regression models—"Baseline 1" (clinical risk factors) and "Baseline 2" (adding BMD, PRS, and grip strength to the Baseline 1 model)—that emulate the variable structure of FRAX (see External Evaluation subsection). These surrogate models served as cohort-specific benchmarks to quantify the incremental value of the longitudinal HyTrax embeddings beyond standard cross-sectional risk factors.

### Model Training and Calibration

#### Training Procedure

To rigorously evaluate model performance and prevent data leakage, participants were split using stratified random sampling into a fixed 70% development set for training/calibration and a held-out 30% validation set for final performance reporting (**Figure 1D**). The 30% held-out validation set contained 8,254 participants, of whom 960 experienced a MOF event and 719 experienced a hip fracture event during follow-up.

These event counts reflect the individualized t₀ design, in which the outcome window begins at each participant’s final recorded visit rather than at a fixed baseline, and may therefore differ from a simple proportional split of the full-cohort event totals. All data preprocessing, including standardization and imputation models, was fitted exclusively on the 70% development split. Within this development set, we employed a nested approach: 10% was set aside for calibration and hyperparameter tuning, while the remaining data were used for model optimization via AdamW with early stopping based on internal validation AUC. Hyperparameters, including latent dimension, number of heads, depth, and dropout, were selected using 5-fold cross-validation performed strictly within the development set (**Table S4**). Final performance metrics were computed solely on the unseen 30% held-out validation set, ensuring an unbiased assessment of generalization capability.

#### Calibration

Post-hoc calibration used Platt scaling implemented as logistic regression on raw model logits to align predicted probabilities with observed event rates ^36^. To ensure equitable performance across diverse populations, we implemented a hierarchical race-aware calibration procedure. Separate Platt scaling models were fitted for each racial/ethnic subgroup that met minimum data sufficiency criteria in the calibration set (N ≥ 20 participants and the presence of at least one outcome event). A global calibration model was fitted on the entire cohort to serve as a robust baseline.

During prediction, individuals belonging to groups with sufficient calibration data were assigned probabilities using their race-specific model. Individuals from underrepresented groups falling below the sufficiency threshold were assigned probability using the global fallback model. In the external FHS evaluation cohort, which consisted exclusively of White participants, the global calibration model was applied to maximize transportability.

### Model Evaluation

Model performance was comprehensively evaluated on the independent WHI validation set and externally evaluated in the FHS cohort. FRAX probabilities were calculated at baseline enrollment and were not recalculated at the individualized HyTrax t₀. This design reflects a pragmatic comparison in which HyTrax uses all longitudinal data available up to t₀ while FRAX represents a clinically standard baseline-only assessment. Accordingly, comparisons between HyTrax and FRAX should be interpreted as reflecting not only differences in model architecture but also differences in information available at prediction time.

#### Standard Metrics and Thresholds

To properly account for censoring, discrimination was assessed using time-dependent AUC at the prespecified 10-year horizon and the IPCW C-index. Calibration was evaluated using calibration-in-the-large, calibration slope, and horizon-specific calibration plots; the Brier score at the 10-year horizon was computed with IPCW. Uncertainty (95% confidence intervals) and between-model comparisons were obtained via nonparametric bootstrap (1,000 replicates); standard ROC metrics and DeLong tests were not utilized.

To ensure consistent application of clinical guidelines, binary classification metrics were calculated by applying the fixed intervention thresholds established by the National Osteoporosis Foundation (NOF): a 10-year probability ≥ 20% for MOF and ≥ 3% for Hip Fracture ^37^. We applied NOF thresholds to all models as practical decision thresholds anchored to clinical practice, recognizing that they were originally defined for FRAX 10-year probabilities. To quantify the incremental improvement of HyTrax over baseline, we calculated the net reclassification improvement (NRI). To assess performance variability in the external evaluation cohort, 95% confidence intervals for the AUC in the FHS dataset were estimated using 1,000 bootstrap replicates.

#### Clinical Utility Analysis

To evaluate the potential impact on clinical decision-making, we performed a decision curve analysis (DCA) to compare the net benefit of the HyTrax ensemble model against standard FRAX baselines across a range of clinically relevant risk thresholds. Additionally, we constructed clinical reclassification tables to quantify the number of fracture cases and non-cases correctly reclassified into appropriate treatment categories based on the FRAX-predicted 10-year probability (e.g., <10%, 10–19%, ≥ 20% for MOF; <3%, ≥ 3% for Hip Fracture) ^38,39^.

### External Evaluation

The HyTrax model trained in WHI was externally evaluated using the original cohort of the FHS data, which provides longitudinal BMD measurements but does not include the variables required to compute FRAX probabilities. Because standard FRAX and FRAX+BMD scores could not be generated from the available dbGaP FHS dataset, we constructed alternative comparator models that approximate the information content of FRAX clinical risk factors and extend beyond them. These comparison models parallel our prior methodological framework ^20^ while being fully aligned with the present HyTrax-based analysis.

#### Baseline Comparator Models in FHS

To enable a meaningful external comparison in the FHS cohort, where standard FRAX scores could not be directly computed, we developed two logistic regression baseline models to emulate the tool’s predictive role (**Table S5**). The first, Baseline 1 (FRAX-CRF Surrogate), was fitted using the available FRAX clinical risk factors present in the FHS dataset: age, sex, height, weight, smoking status, alcohol intake, previous fragility fracture, rheumatoid arthritis, and glucocorticoid use. While parental hip fracture history was utilized during model training and internal validation in the WHI cohort, it was omitted from the FHS baselines due to its unavailability in the dbGaP FHS dataset. This Baseline 1 model served as the closest analogue to the standard FRAX algorithm without BMD. The second, Baseline 2 (Extended Clinical + BMD Model), incorporated these core risk factors along with total hip and lumbar spine BMD, PRS, and grip strength. Because grip strength was sparsely measured at the baseline examination in FHS (N=17), this feature relied heavily on imputed placeholder values in the external evaluation, acting as a sparse predictor rather than a routine clinical measurement to maintain structural consistency with the models derived in WHI. This extended model reflects the maximum predictive information available in the external cohort, providing a more stringent benchmark for evaluating the incremental value of HyTrax. Because Baseline 1 and 2 were fitted internally within the FHS cohort to provide a contemporary localized benchmark, this analysis serves as an evaluation of the HyTrax embeddings’ transportability rather than a strict external evaluation of a fully locked clinical algorithm.

#### Hybrid Comparison Model in FHS

To validate the hybrid strategy in the external cohort, we applied the fixed ensemble weighting optimized in the WHI development phase. Specifically, the longitudinal risk score generated by HyTrax for each FHS participant was combined with the risk probability from the Baseline 1 (FRAX-CRF Surrogate) and Baseline 2 (Extended Clinical + BMD Model) models using the fixed α coefficient derived from WHI.

While the HyTrax model weights and hybrid integration parameters (α) were strictly fixed from the WHI development phase to test transportability, the baseline clinical comparator models (Baseline 1 and 2) were estimated directly within the FHS cohort to provide localized standard-of-care benchmarks.

### Explainability and Model Interpretation

Model interpretability was assessed using SHapley Additive exPlanations (SHAP) values ^40^. SHAP summary plots quantified the global contributions of both static baseline factors and longitudinal features. To facilitate clinical interpretation, dynamic predictors are presented using descriptive labels. These labels correspond to the underlying tokenized longitudinal inputs provided to the Transformer encoder, which compute absolute measurements at specific visits, raw visit-to-visit differences, and normalized rates of change. These token-level dynamic features allow the model to learn subtle temporal patterns in physical decline that are not captured by static predictors alone.

### Implementation and Ethical Approval

All deep-learning components were implemented in PyTorch v2.3 and executed on NVIDIA A100 GPUs at the Ohio Supercomputer Center ^41^. Mixed-effects models were fit using “statsmodels” ^42^. To rigorously account for censoring without relying on unvalidated custom routines, all time-dependent evaluations, including the IPCW C-index, time-dependent AUC, and horizon-specific Brier scores, were conducted using the scikit-survival and lifelines Python libraries ^43^. Multiple imputation was performed using the mice package in R ^29^.

All data analyses were conducted in accordance with approved institutional protocols. The WHI and FHS are federally regulated cohort studies in which all participants provided written informed consent at enrollment. Use of WHI and FHS data for the present research was approved by the institutional review board (IRB) at The Ohio State University (protocol STUDY20250826). Access to controlled-use data was granted through dbGaP data use agreements, and all analyses complied with relevant data-use and privacy regulations.

## Results

### Study Population Characteristics

Participants who experienced fracture events differed significantly from those who did not across nearly all baseline clinical characteristics (**Table 1** and **Table S6**). Women who sustained a MOF were significantly older (mean 68.4 vs. 63.1 years, P<0.001), had lower body weight (71.3 vs. 77.4 kg, P<0.001), and demonstrated lower BMD at the hip (0.7 vs. 0.9 g/cm², P<0.001) compared to fracture-free participants.

**Table 1.** Comparison of Baseline Demographic and Clinical Characteristics of Women’s Health Initiative Participants, Stratified by Major Osteoporotic Fractures (MOF) Event (N=27,512).

| Variable | Participants without MOF<br>(N=24,915) | Participants with MOF<br>(N=2,597) | P-value* |
| --- | --- | --- | --- |
| Age (years), mean (SD) | 63.1 (7.4) | 68.4 (6.7) | <0.001 |
| Height (cm), mean (SD) | 161 (6.3) | 162 (6.6) | <0.001 |
| Weight (kg), mean (SD) | 77.4 (17.1) | 71.3 (14.9) | <0.001 |
| FRAX (without BMD) for MOF, mean (SD) | 7.3 (6.6) | 15.3 (9.1) | <0.001 |
| FRAX (with BMD) for MOF, mean (SD) | 7.1 (6.0) | 14.9 (8.1) | <0.001 |
| Hip BMD (g/cm<sup>2</sup>), mean (SD) | 0.9 (0.2) | 0.7 (0.1) | <0.001 |
| Spine BMD (g/cm<sup>2</sup>), mean (SD) | 1.0 (0.1) | 0.9 (0.1) | <0.001 |
| Grip Strength (kg), mean (SD) | 23.3 (6.2) | 22.4 (6.1) | 0.03 |
| PRS <sup>a</sup>, mean (SD) | 1.2 (25.8) | 7.2 (23.1) | <0.001 |
| RACE/ethnicity, *n* (%) |  |  | <0.001 |
| American Indian or Alaskan Native | 569 (2.3%) | 22 (0.8%) |  |
| Asian or Pacific Islander | 478 (1.9%) | 21 (0.8%) |  |
| African-American | 9,910 (39.8%) | 145 (5.6%) |  |
| Hispanic/Latino | 4,499 (18.1%) | 135 (5.2%) |  |
| Non-Hispanic White | 9,459 (38.0%) | 2,274 (87.6%) |  |
| Current Smoking, *n* (%) | 2,355 (9.5%) | 224 (8.6%) | 0.07 |
| Rheumatoid Arthritis, *n* (%) | 3,339 (13.4%) | 332 (12.8%) | 0.39 |
| Previous fragility fracture, *n* (%) | 303 (1.2%) | 85 (3.3%) | <0.001 |
| Parental hip fracture history, *n* (%) | 771 (3.1%) | 151 (5.8%) | 0.01 |
| Alcohol intake, *n* (%) | 16,155 (64.8%) | 1,852 (71.3%) | <0.001 |
| Glucocorticoid use, *n* (%) | 39 (0.2%) | 9 (0.3%) | 0.05 |
| Diabetes, *n* (%) | 2,229 (8.9%) | 171 (6.6%) | 0.33 |
| ≥ 1 Previous fall, *n* (%) | 7,904 (31.7%) | 1,060 (40.8%) | <0.001 |

**Abbreviations:** FRAX, Fracture Risk Assessment Tool; SD, Standard Deviation; BMD, Bone Mineral Density; PRS, Polygenic Risk Score; MOF, Major Osteoporotic Fractures.
\* P-value was obtained by t-test for continuous variables and chi-square tests for the categorical variable.
<sup>a</sup> Developed using the LDpred algorithm with 103,155 genetic variants from UK Biobank GWAS summary statistics. Major Osteoporotic Fracture was defined as a fracture occurring at the hip, spine, shoulder, or wrist.

These trends were even more pronounced for hip fracture cases, where the mean FRAX probability (without BMD) was nearly triple that of non-cases (5.6% vs. 1.9%, P<0.001). Clinical risk factors such as a history of previous falls and prior fragility fractures were also significantly more prevalent in the fracture groups. As detailed in **Table S3**, while anthropometric measurements (height and weight) were dense across visits, repeated measures for specific musculoskeletal traits, such as hip BMD and grip strength, were available only for subsets of participants, meaning trajectory information for those specific modalities relied on missingness indicators for some individuals.

The external evaluation cohort (FHS, N=1,193) presented a distinct clinical profile compared to the WHI dataset (**Table S7**). FHS participants were significantly older (mean 75.3 vs. 63.7 years, P<0.001) and had lower mean weight (69.9 vs. 76.7 kg, P<0.001) and hip BMD (0.7 vs. 0.9 g/cm², P<0.001). While the WHI cohort was racially diverse, the FHS sample was exclusively Non-Hispanic White (100%). Reflecting their advanced age and lower bone density, the FHS cohort had a higher prevalence of fracture outcomes: 14.5% experienced an MOF and 10.1% experienced a hip fracture during follow-up, compared with 9.4% and 6.5% in the full WHI cohort, respectively.

### Model Discrimination

On the WHI internal validation set (N=8,254), the HyTrax + FRAX (BMD) model achieved the higher discrimination in this analysis, with the time-dependent AUC of 0.85 (95% CI 0.84–0.86) (**Figure 2**). The HyTrax model alone, which integrated tokenized longitudinal trajectories of BMD, grip strength, height, and weight with subject-specific mixed-effects slopes and static clinical covariates, without incorporating FRAX probabilities, achieved the time-dependent AUC of 0.80. While the standalone HyTrax model performed comparably to the baseline FRAX (BMD) model but did not exceed it, the integration of these distinct signal sources in the HyTrax + FRAX (BMD) ensemble model outperformed the FRAX (BMD) baseline (P<0.001). The time-dependent ROC and PR curves illustrate this additive value, showing clear separation for the hybrid ensemble and improved precision under class imbalance.

**Figure 2.**
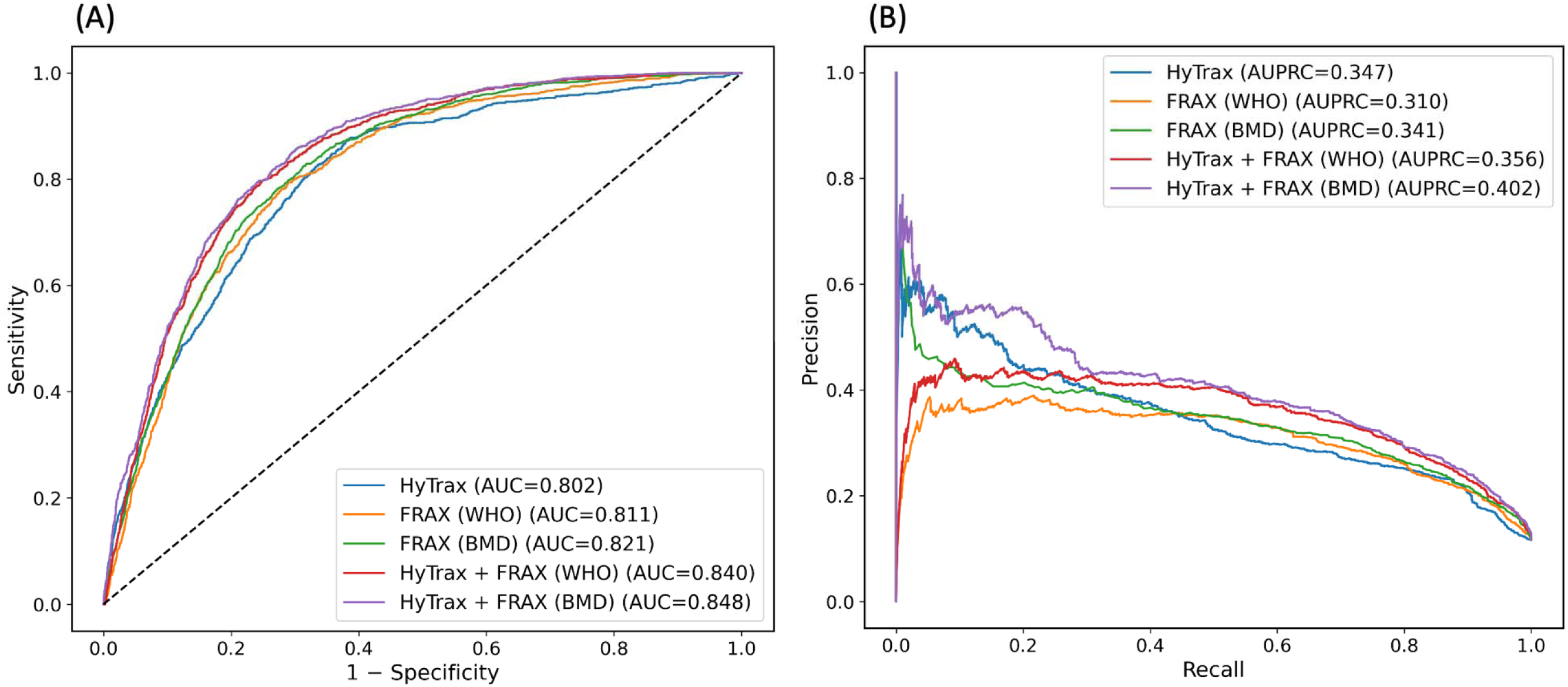
Time-Dependent Discrimination Performance of the Hybrid Trajectory-Based (HyTrax) Model for Major Osteoporotic Fracture (MOF) at a 10-Year Horizon. Panels summarize the internal discrimination performance of the predictive models on the held-out WHI validation set (N = 8,254). Curves are shown for the Fracture Risk Assessment Tool (FRAX) (WHO) (clinical risk factors only), FRAX (BMD) (clinical factors + femoral neck BMD), HyTrax alone, and the HyTrax + FRAX (BMD) hybrid ensemble. (A) Time-dependent Receiver Operating Characteristic (ROC) curves illustrate the trade-off between sensitivity and specificity across decision thresholds. The hybrid ensemble demonstrates the higher time-dependent Area Under the Curve (AUC = 0.85) in this analysis, outperforming both the standard FRAX baselines and the standalone HyTrax model. (B) Precision–Recall (PR) curves evaluate model performance under class imbalance by depicting precision across levels of recall, indicating improved precision across recall levels.

These findings were replicated in the FHS external evaluation cohort, where HyTrax again outperformed FRAX-like logistic regression baselines constructed from available clinical variables (**Table 2**). Despite differences in racial composition, measurement intervals, and fracture adjudication procedures between the two cohorts, the longitudinal Transformer architecture exhibited stable predictive behavior, suggesting transportability of the longitudinal sequence representations.

**Table 2.**
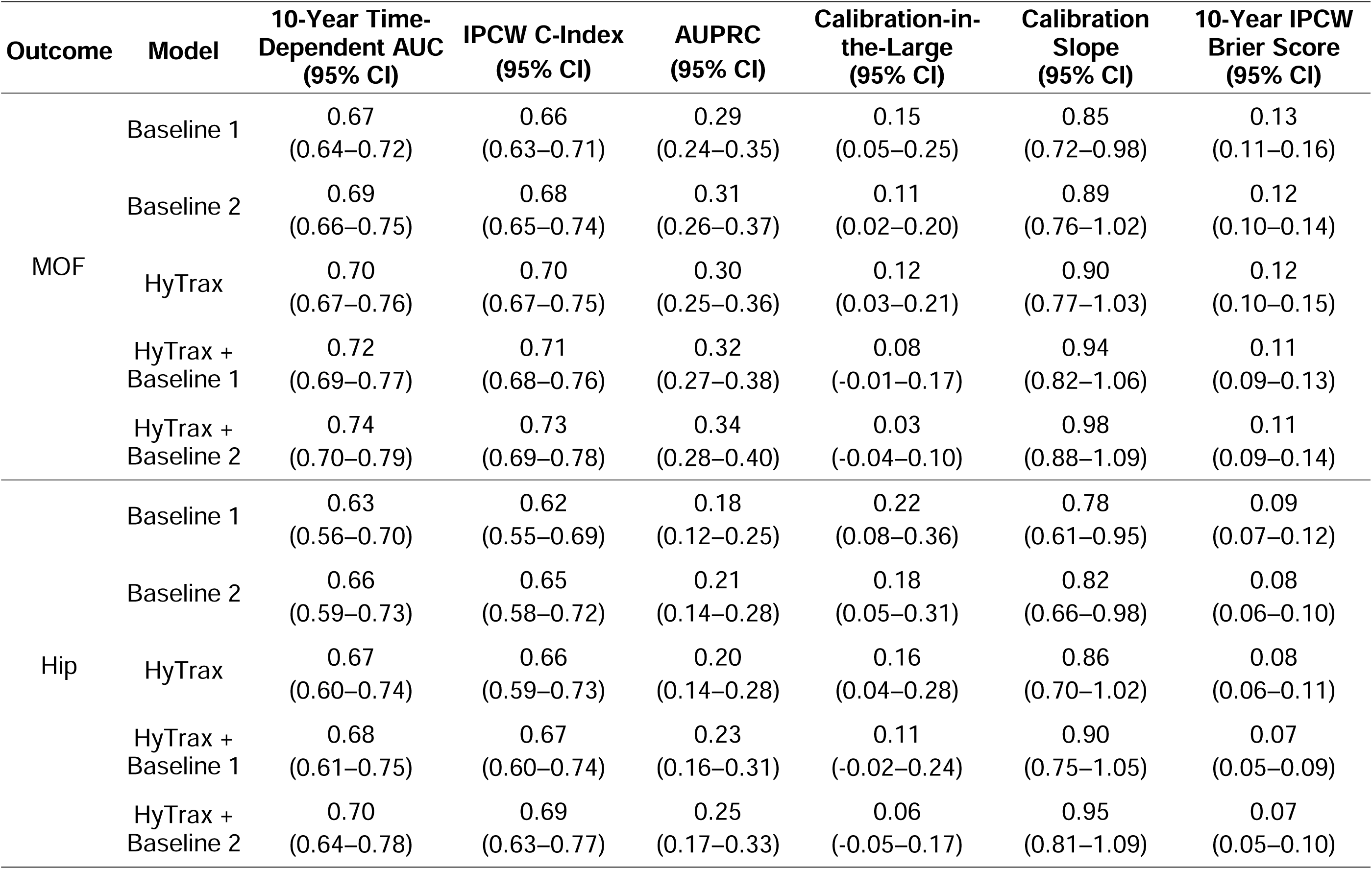
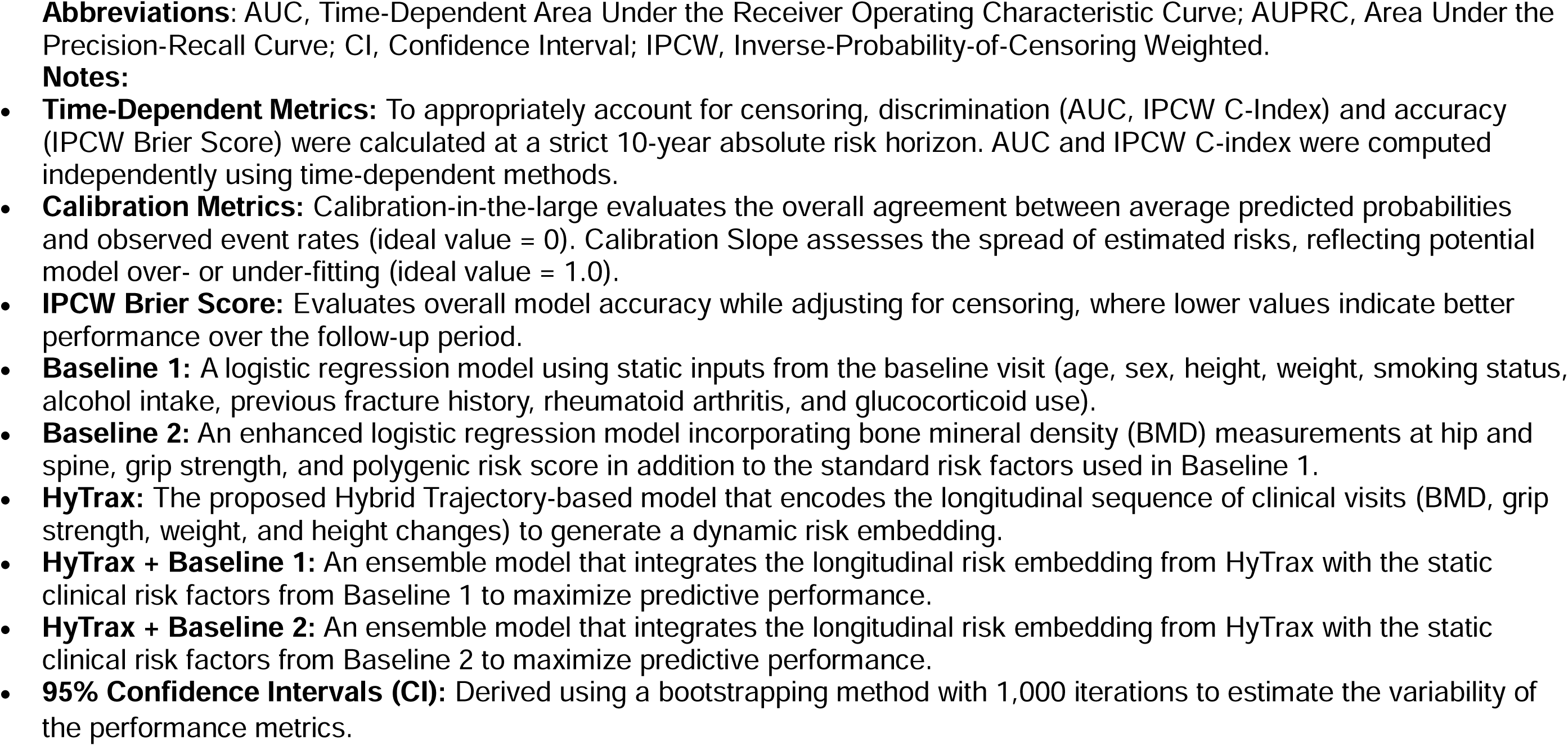
Performance of the Hybrid Trajectory-Based (HyTrax) Model and Baseline Models in the Independent Framingham Heart Study (FHS) Evaluation Cohort (N=1,193) for Major Osteoporotic Fractures (MOF) and Hip Fracture at a 10-Year Horizon. .

| Outcome | Model | 10-Year Time-Dependent AUC<br>(95% CI) | IPCW C-Index<br>(95% CI) | AUPRC<br>(95% CI) | Calibration-in-the-Large<br>(95% CI) | Calibration Slope<br>(95% CI) | 10-Year IPCW Brier Score<br>(95% CI) |
| --- | --- | --- | --- | --- | --- | --- | --- |
| MOF | Baseline 1 | 0.67<br>(0.64–0.72) | 0.66<br>(0.63–0.71) | 0.29<br>(0.24–0.35) | 0.15<br>(0.05–0.25) | 0.85<br>(0.72–0.98) | 0.13<br>(0.11–0.16) |
|  | Baseline 2 | 0.69<br>(0.66–0.75) | 0.68<br>(0.65–0.74) | 0.31<br>(0.26–0.37) | 0.11<br>(0.02–0.20) | 0.89<br>(0.76–1.02) | 0.12<br>(0.10–0.14) |
|  | HyTrax | 0.70<br>(0.67–0.76) | 0.70<br>(0.67–0.75) | 0.30<br>(0.25–0.36) | 0.12<br>(0.03–0.21) | 0.90<br>(0.77–1.03) | 0.12<br>(0.10–0.15) |
|  | HyTrax + Baseline 1 | 0.72<br>(0.69–0.77) | 0.71<br>(0.68–0.76) | 0.32<br>(0.27–0.38) | 0.08<br>(-0.01–0.17) | 0.94<br>(0.82–1.06) | 0.11<br>(0.09–0.13) |
|  | HyTrax + Baseline 2 | 0.74<br>(0.70–0.79) | 0.73<br>(0.69–0.78) | 0.34<br>(0.28–0.40) | 0.03<br>(-0.04–0.10) | 0.98<br>(0.88–1.09) | 0.11<br>(0.09–0.14) |
| Hip | Baseline 1 | 0.63<br>(0.56–0.70) | 0.62<br>(0.55–0.69) | 0.18<br>(0.12–0.25) | 0.22<br>(0.08–0.36) | 0.78<br>(0.61–0.95) | 0.09<br>(0.07–0.12) |
|  | Baseline 2 | 0.66<br>(0.59–0.73) | 0.65<br>(0.58–0.72) | 0.21<br>(0.14–0.28) | 0.18<br>(0.05–0.31) | 0.82<br>(0.66–0.98) | 0.08<br>(0.06–0.10) |
|  | HyTrax | 0.67<br>(0.60–0.74) | 0.66<br>(0.59–0.73) | 0.20<br>(0.14–0.28) | 0.16<br>(0.04–0.28) | 0.86<br>(0.70–1.02) | 0.08<br>(0.06–0.11) |
|  | HyTrax + Baseline 1 | 0.68<br>(0.61–0.75) | 0.67<br>(0.60–0.74) | 0.23<br>(0.16–0.31) | 0.11<br>(-0.02–0.24) | 0.90<br>(0.75–1.05) | 0.07<br>(0.05–0.09) |
|  | HyTrax + Baseline 2 | 0.70<br>(0.64–0.78) | 0.69<br>(0.63–0.77) | 0.25<br>(0.17–0.33) | 0.06<br>(-0.05–0.17) | 0.95<br>(0.81–1.09) | 0.07<br>(0.05–0.10) |

**Abbreviations:** AUC, Time-Dependent Area Under the Receiver Operating Characteristic Curve; AUPRC, Area Under the Precision-Recall Curve; CI, Confidence Interval; IPCW, Inverse-Probability-of-Censoring Weighted.
**Notes:**

### Clinical Utility and Net Benefit

Decision curve analysis demonstrated that the HyTrax ensemble model provided a higher net benefit than both FRAX baselines across the standard intervention thresholds for MOF and Hip Fracture (**Figure 3**). Reclassification analysis further detailed these shifts in risk categorization **(Tables 3-4)**. For MOF, among women who eventually experienced a fracture, the hybrid model reclassified 54.7% (525/960) into a higher risk category compared to FRAX (BMD) alone (**Table 3**). This increased sensitivity was accompanied by a trade-off, as 28.2% (2,055/7,294) of non-fracture cases were also reclassified upward. However, decision curve analysis (**Figure 3**) indicated that the overall net benefit of the hybrid model remained positive and exceeded that of both FRAX baselines across the relevant 10–20% decision thresholds, yielding an overall NRI of +26.5% relative to the standard FRAX (BMD) model. A similar reclassification pattern was observed for Hip Fracture risk. The hybrid model reclassified 29.2% (210/719) of fracture cases upward into the treatment range (≥ 3%). Concurrently, it down-classified 45.4% (3,419/7,535) of non-cases into the low-risk category (<3%) (**Table 4**).

**Figure 3.**
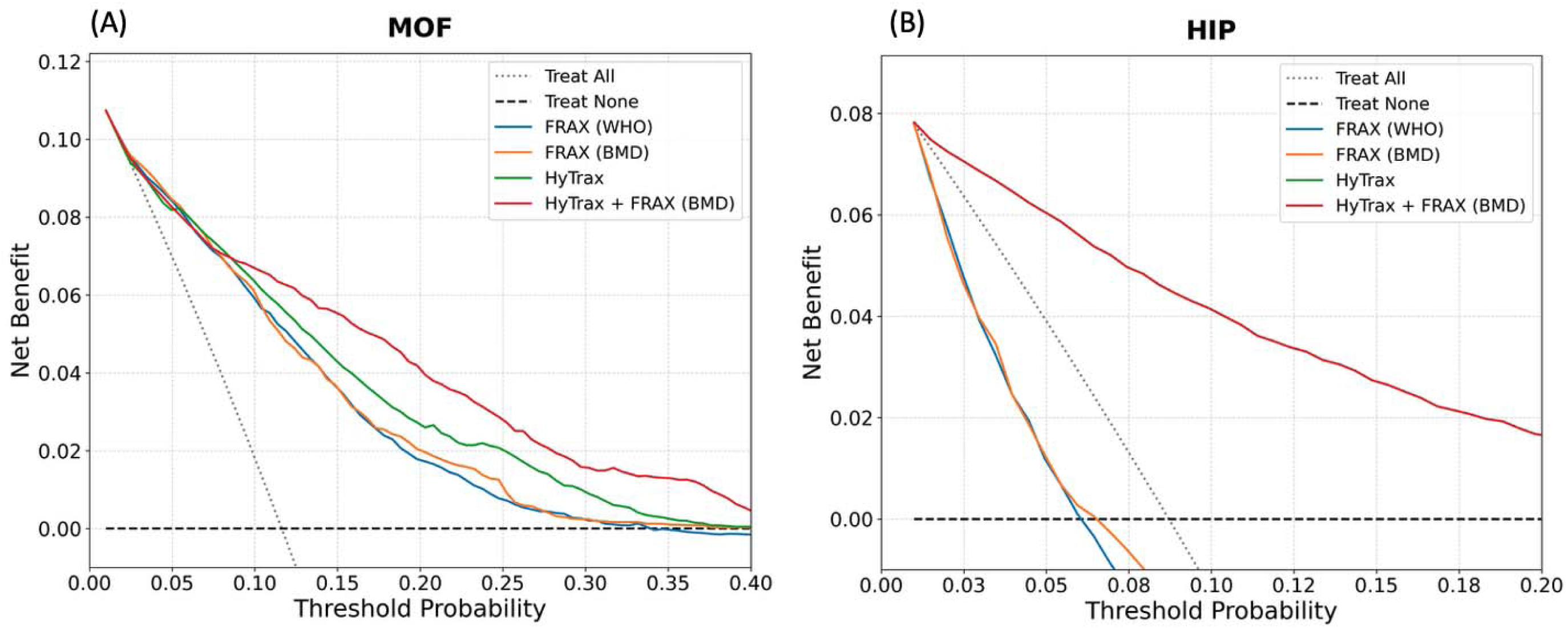
Decision Curve Analysis of the Hybrid Trajectory-Based (HyTrax) Model Compared to the Standard Fracture Risk Assessment Tool (FRAX) Baselines. Plots illustrate the net benefit of the HyTrax hybrid model (red line) versus standard FRAX clinical (blue) and FRAX-BMD (orange) models across a range of decision thresholds. (A) Major Osteoporotic Fracture (MOF): The hybrid model shows improved net benefit across standard intervention thresholds (10–20%). (B) Hip Fracture: The hybrid model maintains higher net benefit specifically around the 3% treatment threshold used in clinical guidelines. The "Treat All" (gray) and "Treat None" (black) lines serve as reference strategies. All curves were generated using the WHI internal validation set (N = 8,254).

**Table 3.**
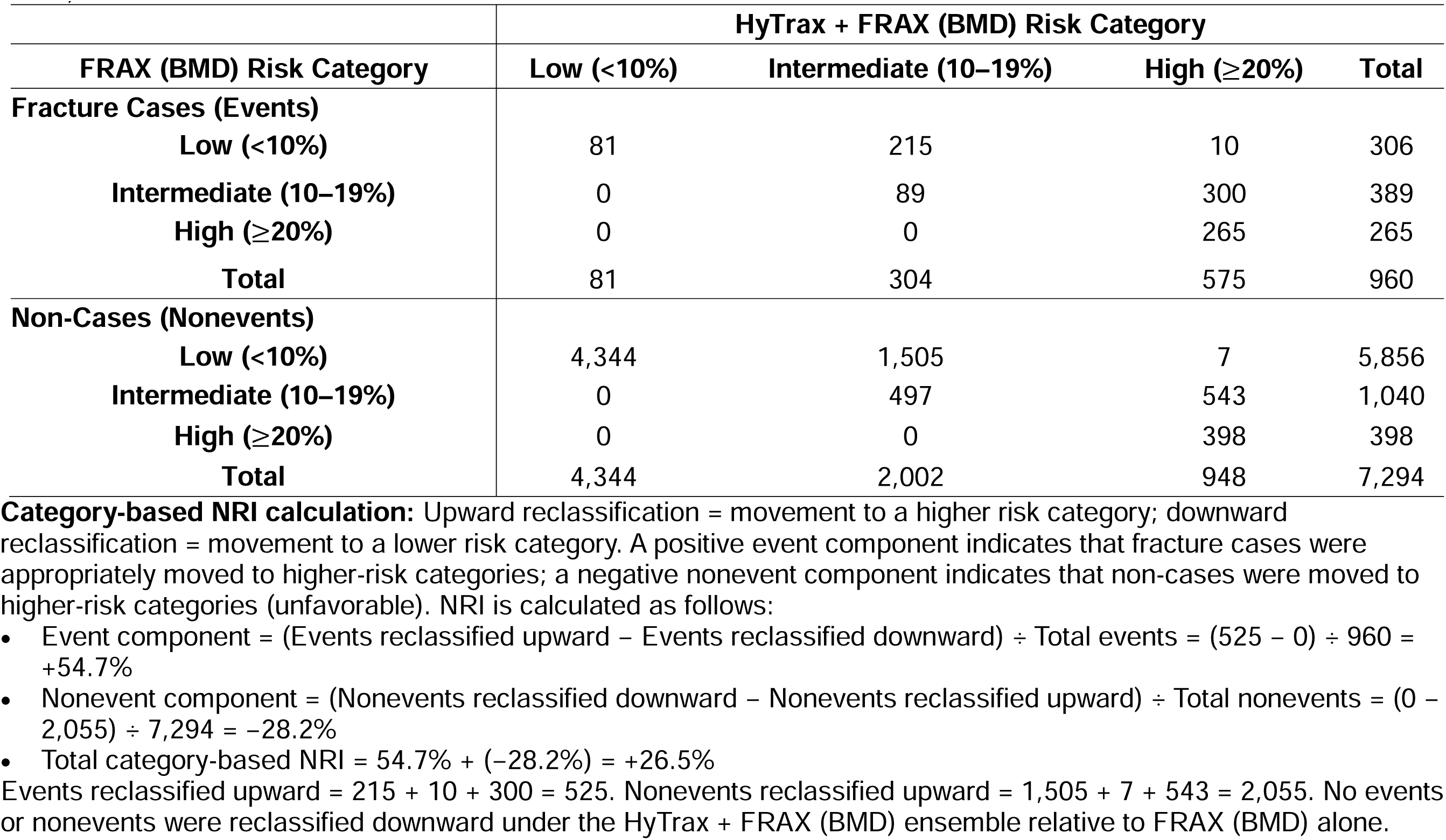
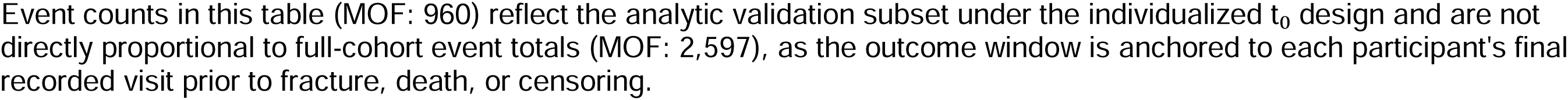
Clinical Reclassification of Major Osteoporotic Fracture (MOF) Risk Prediction by the HyTrax + FRAX (BMD) Ensemble Compared to FRAX (BMD) Alone. This table shows the movement of participants across clinical risk categories when switching from the standard FRAX (BMD) model to the hybrid ensemble HyTrax + FRAX (BMD) model. Risk categories are defined based on National Osteoporosis Foundation (NOF) intervention thresholds: Low (<10%), Intermediate (10–19%), and High ( 20%) for MOF. All results were generated using the WHI internal validation set (N = 8,254).

**Table 4.** Clinical Reclassification of Hip Fracture Risk Prediction by the HyTrax + FRAX (BMD) Ensemble Compared to FRAX (BMD) Alone. This table shows the movement of participants across clinical risk categories when switching from the standard FRAX (BMD) model to the hybrid ensemble. Risk categories are defined based on National Osteoporosis Foundation (NOF) intervention thresholds: Low (<3%) and High ( 3%) for Hip Fracture. All results were generated using the WHI internal validation set (N = 8,254).

| FRAX (BMD) Risk Category | HyTrax + FRAX (BMD) Risk Category |  |  |
| --- | --- | --- | --- |
| | Low (<3%) | High ($\geq 3\%$) | Total |
| **Fracture Cases (Events)** |  |  |  |
| Low (<3%) | 19 | 210 | 229 |
| High ($\geq 3\%$) | 45 | 445 | 490 |
| **Total** | 64 | 655 | 719 |
| **Non-Cases (Nonevents)** |  |  |  |
| Low (<3%) | 1,235 | 922 | 2,157 |
| High ($\geq 3\%$) | 3,419 | 1,959 | 5,378 |
| **Total** | 4,654 | 2,881 | 7,535 |

**Direction of reclassification:** A participant is reclassified upward if HyTrax + FRAX (BMD) assigns them to High ( $\geq 3\%$ ) when FRAX (BMD) alone assigned them to Low (<3%); reclassified downward if assigned to Low when FRAX (BMD) assigned High. Upward reclassification is favorable for fracture cases (correctly identified as high-risk) and unfavorable for non-cases (incorrectly flagged as high-risk).
**Category-based NRI components** (reported for transparency; hip NRI is not included as a primary endpoint in the main analysis):
- Event component = $(210 - 45) \div 719 = 165 \div 719 = +22.9\%$ (net upward reclassification among hip fracture cases) - Nonevent component = $(3,419 - 922) \div 7,535 = 2,497 \div 7,535 = +33.1\%$ (net downward reclassification among non-cases, favorable) - Total category-based NRI = $22.9\% + 33.1\% = +56.1\%$
Event counts in this table (Hip fracture: 719) reflect the analytic validation subset under the individualized $t_0$ design and are not directly proportional to full-cohort event totals (Hip fracture: 1,797), as the outcome window is anchored to each participant's final recorded visit prior to fracture, death, or censoring.

### Hybrid Model Behavior

The hybrid estimator, P_hybrid_=α P_HyTrax_+(1-α) P_FRAX_, dynamically weighted the HyTrax and FRAX contributions. Optimization on the WHI calibration subset yielded α=0.15 for the FRAX-only comparison and α=0.20 for the FRAX-BMD comparison. These values indicate that FRAX remains the dominant global signal, but even a 20% contribution from longitudinal deep learning materially improves accuracy.

### Explainability and Feature Importance

SHAP-based explainability revealed that specific dynamic predictors, most notably longitudinal weight fluctuations and overall height loss trajectories, had a profound influence on model predictions alongside static baseline factors **(Figures 4** **& S3)**. The largest overall contributors to predicted fracture risk included baseline age, longitudinal weight dynamics, baseline height, and PRS. Notably, the PRS emerged as a key static predictor, ranking as the sixth most important feature overall, reinforcing the value of genetic predisposition alongside phenotypic trajectories. While its contribution was lower than the longitudinal dynamic features (BMD/grip slopes), it consistently ranked higher than most traditional clinical risk factors (e.g., smoking, alcohol), demonstrating the value of intrinsic genetic susceptibility alongside phenotypic trajectories. Token-level dynamic features provided granular signals that allowed the HyTrax to detect subtle deterioration patterns. Static values alone contributed less, reinforcing the importance of modeling physiologic change rather than relying solely on baseline status.

**Figure 4.**
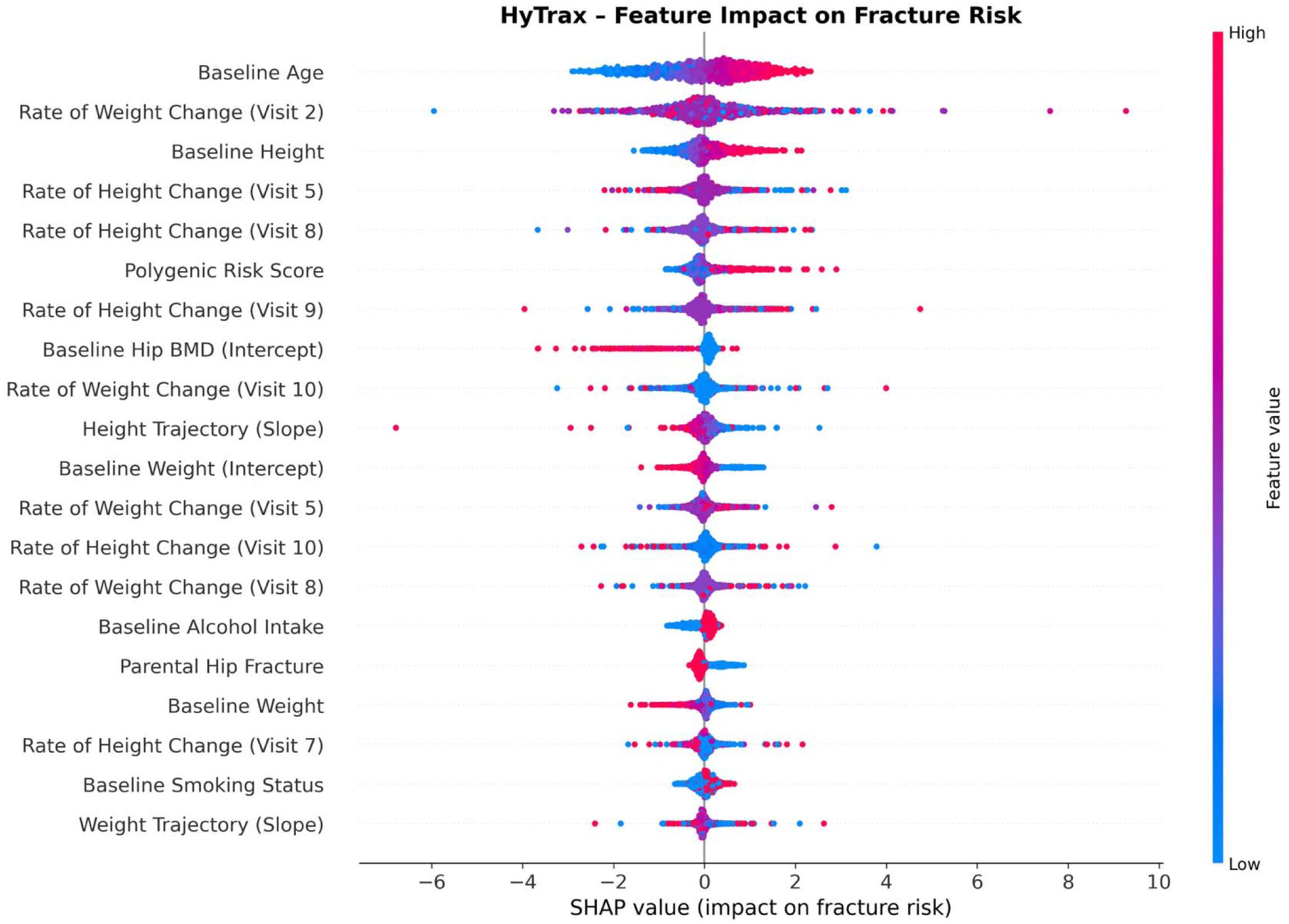
Global Feature Importance of the Hybrid Trajectory-Based (HyTrax) Model Using SHapley Additive exPlanations (SHAP) Values. The beeswarm plot shows the contribution of each feature to the predicted fracture risk across all individuals. Features are ranked from top to bottom by their mean absolute SHAP value on the held-out WHI internal validation set (N=8,254), with colors indicating the underlying feature magnitude (blue = low, red = high).

### External Evaluation in FHS

For MOF, the standalone HyTrax model (time-dependent AUC 0.70, 95% CI 0.67–0.76) outperformed the clinical risk factor model (Baseline 1, AUC 0.67, 95% CI 0.64-0.72) and achieved discrimination comparable to the BMD-enhanced Baseline 2 (AUC 0.69, 95% CI 0.66-0.75). The HyTrax + Baseline 2 ensemble achieved the higher discrimination (AUC 0.74, 95% CI 0.70-0.79) and precision (AUPRC 0.34, 95% CI 0.28-0.40) while maintaining the more favorable calibration metrics (Calibration-in-the-Large 0.03, 95% CI -0.04-0.10), confirming that longitudinal embeddings capture prognostic signals orthogonal to static bone density (**Table 2**).

For Hip Fracture, the performance gains were similarly evident, though overall discrimination was lower than for MOF, reflecting the smaller number of events (N=120) and the challenge of long-term prediction in this older cohort. The ensemble of HyTrax + Baseline 2 achieved an AUC of 0.70 (95% CI 0.64–0.78), outperforming Baseline 1 (AUC 0.63, 95% CI 0.56-0.70) and providing a meaningful gain over Baseline 2 (AUC 0.66, 95% CI 0.59-0.73). Despite the wider confidence intervals inherent to the smaller sample size, the ensemble maintained stable calibration (Brier Score 0.07, 95% CI 0.05-0.10).

## Discussion

### Integrating Temporal Dynamics with Established Clinical Risk Tools

In this study, we establish that combining individualized musculoskeletal trajectories with standard clinical tools fundamentally improves fracture prediction. By developing the Transformer-based HyTrax framework, we demonstrate that the HyTrax + FRAX (BMD) ensemble successfully merges two complementary predictive paradigms: the population-anchored stability of traditional FRAX and the personalized, temporal dynamism of deep sequential learning, yielding improved gains in accuracy, calibration, and clinical utility. While FRAX provides a robust baseline anchored in decades of epidemiologic evidence, it treats patients as static data points at a single moment in time. By contrast, our results show that the trajectory of physiological change, specifically the subject-specific rates of bone density loss, muscle weakening, height and weight change, carries independent prognostic information that static baselines miss.

This finding aligns with recent longitudinal studies suggesting that the velocity of bone loss is a more potent predictor of fracture than absolute mass alone ^44–47^. For instance, Kim et al. demonstrated that rapid bone loss is inextricably linked to simultaneous muscle deterioration, creating a "resorption-dominant" phenotype that precedes clinical fractures ^8^. By capturing these coupled, non-linear trajectories using LMM slopes and attention-based sequence modeling, HyTrax effectively identifies high-risk phenotypes that may not yet meet the osteoporotic threshold (T-score ≤ -2.5) but are nonetheless rapidly deteriorating. Furthermore, the high feature importance ranking of the PRS confirms that genetic predisposition provides a stable, complementary risk signal that operates independently of these dynamic musculoskeletal changes.

### Synergy Between Transformer Dynamics and FRAX Clinical Knowledge

The hybrid integration framework capitalized on the complementary strengths of the Transformer model and FRAX. FRAX provides clinically validated, population-level risk estimates anchored in decades of epidemiologic evidence ^38,48^, but it lacks the ability to incorporate within-person longitudinal change. Conversely, the Transformer excels at modeling nonlinear dependencies and individualized temporal patterns but risks overfitting without anchoring to population priors. The hybrid probability estimator, using an optimized weighting coefficient α ≈ 0.15–0.20, balanced these contributions. A small α indicates that FRAX establishes the absolute risk landscape, while the Transformer refines patient-level ranking based on bone, muscle, weight, and height trajectories. This synergy produced meaningful gains in discrimination, calibration, and reclassification, including an NRI of +26.5% relative to FRAX-BMD. We applied the NOF intervention thresholds (20% MOF, 3% hip fracture) to all models as practical decision thresholds anchored to clinical practice, recognizing that while they were originally defined for FRAX 10-year probabilities, they serve as the most relevant benchmark for evaluating the potential clinical utility of new predictive tools.

### Mechanistic Insights into Temporal Learning

The observed performance gains likely reflect the Transformer’s capacity to detect subtle and early “micro-patterns” in physiological decline. Unlike static models that rely on single measurements or simple two-point changes, the Transformer distributes attention across multiple visits, enabling it to capture nonlinear acceleration in BMD loss, progressive decline in grip strength, and coordinated changes across musculoskeletal systems. Despite reliance on missingness indicators for participants with sparse repeated measures, SHAP analysis indicated that available temporal features such as weight and height dynamics, contributed to the risk predictions, aligning with understood pathophysiological patterns ^8–12^. The model’s sensitivity to evolving longitudinal patterns, rather than absolute values alone, suggests that deep-learning–based trajectory modeling may offer a clinically meaningful window for early intervention.

### Advancing Equity Through Race-Aware Calibration

Fracture risk prediction tools have historically exhibited bias, often overestimating or underestimating risk in non-White populations due to derivation cohorts that were predominantly of European descent ^14^. We addressed this by implementing a race-aware calibration strategy that prioritizes subgroup specificity while maintaining stability. By fitting distinct mapping functions for racial groups with sufficient sample size (e.g., Black, Hispanic, and White women in WHI) and using a global fallback for smaller groups, we aimed to mitigate prediction disparities. This approach ensures that risk estimates are tailored to the observed fracture rates within specific subpopulations where data allows, without overfitting to sparse groups. These findings highlight that while deep learning models can learn universal physiological signals, their translation into absolute risk probabilities benefits from population-specific post-processing.

### Clinical Implications and Future Deployment

The HyTrax model provides a framework for dynamic, personalized risk assessment that integrates both temporal patterns of physiological change and well-established clinical risk factors. This approach could be deployed as a web-based or electronic health record, embedded tool, allowing clinicians to update risk predictions as new visits become available. Such a system could enable earlier identification of high-risk trajectories, support shared decision-making regarding osteoporosis treatment, and provide transparent, explainable outputs through SHAP-derived importance metrics. By bridging the gap between population-level risk tools and personalized longitudinal modeling, HyTrax represents a step toward precision fracture prevention.

### Limitations and Future Directions

This study has several limitations. The WHI and FHS cohorts lack certain clinical, behavioral, and biochemical markers relevant to fracture risk, and differences in visit spacing and DXA instrumentation introduce cohort-specific variability. External evaluation was limited to the predominantly White FHS cohort, restricting assessment of racial generalizability. Although osteoporosis medication users were excluded at baseline, participants initiating therapy during follow-up were not censored; this treatment paradox may cause the model to underestimate true risk in successfully treated high-risk individuals. Additionally, FRAX probabilities were derived at baseline rather than at the individualized HyTrax prediction anchor (t₀), so performance comparisons reflect both architectural differences and information asymmetry. Finally, as shown in **Table S3**, hip BMD and grip strength measurements were sparse for many participants, requiring the model to rely on static features and missingness indicators for those individuals; sensitivity analyses in cohorts with complete multi-visit trajectories are needed to isolate the predictive contribution of longitudinal tokens.

Future work should expand HyTrax to incorporate additional biomarkers such as bone turnover markers and imaging-derived microarchitecture, extend evaluation to men and younger adults, and incorporate time-varying treatment as a censoring event to better isolate the natural history of musculoskeletal decline. Continuous-time Transformers and irregular-interval attention mechanisms may improve modeling of real-world clinical data. Prospective validation and cost-effectiveness analyses will be essential before clinical deployment, and integration with electronic health records could enable dynamically updated risk assessment in routine practice.

## Conclusion

In summary, this study addresses the long-standing clinical uncertainty regarding the value of repeated musculoskeletal assessments by demonstrating that individualized temporal dynamics enhance fracture prediction. While current guidelines and tools such as FRAX rely on static "snapshots," our findings show that the trajectory of bone, muscle, weight, and height loss carries independent prognostic information that baseline measures miss. By synergizing these Transformer-derived longitudinal signals with the robust population priors of FRAX, the HyTrax + FRAX (BMD) ensemble achieves improved discrimination, calibration, and supports further evaluation across cohorts with distinct clinical profiles. Although prospective validation in broader populations is necessary, this framework supports a shift from static risk stratification toward a dynamic, precision prevention model capable of identifying high-risk trajectories before fractures occur.

## Data Availability

The data used in the current study is available through controlled access of the database of Genotype and Phenotype (dbGap) (https://www.ncbi.nlm.nih.gov/projects/gap/cgi-bin/study.cgi?study_id=phs000200.v12.p3). The summary statistics of the Genome-Wide Association Study used in the current study are available from the GEnetic Factors for Osteoporosis Consortium at http://www.gefos.org/?q=content/data-release-2018.
All data utilized for this study were obtained from publicly accessible resources via the Database of Genotypes and Phenotypes (dbGaP). Data used for the discovery phase (Women's Health Initiative, WHI) were sourced from several genetic initiatives, including the WHI Sequencing Project (phs000281) and the WHI Memory Study (phs000675). Additional associated genetic resources leveraged include the Population Architecture using Genomics and Epidemiology (PAGE) study (phs000227), the Genomics and Randomized Trials Network (GARNET) study (phs000315), and the SNP Health Association Resource (SHARe) study (phs000386). The primary cohort used for evaluation, the Framingham Heart Study (FHS), is available under accession number phs000007.

## Author contributions

Conceptualization: Jongyun Jung. Data curation: Jongyun Jung.

Formal analysis: Jongyun Jung. Funding acquisition: Qing Wu. Investigation: Jongyun Jung, Qing Wu. Methodology: Jongyun Jung.

Project administration: Jongyun Jung, Qing Wu. Resources: Qing Wu.

Software: Jongyun Jung. Supervision: Qing Wu.

Validation: Jongyun Jung. Visualization: Jongyun Jung.

Writing –original draft: Jongyun Jung.

Writing –review & editing: Jongyun Jung, Qing Wu.

## Data Sharing Statement

The data used in the current study is available through controlled access of the database of Genotype and Phenotype (dbGap) (https://www.ncbi.nlm.nih.gov/projects/gap/cgi-bin/study.cgi?study_id=phs000200.v12.p3). The summary statistics of the Genome-Wide Association Study used in the current study are available from the GEnetic Factors for Osteoporosis Consortium at http://www.gefos.org/?q=content/data-release-2018

All data utilized for this study were obtained from publicly accessible resources via the Database of Genotypes and Phenotypes (dbGaP). Data used for the discovery phase (Women’s Health Initiative, WHI) were sourced from several genetic initiatives, including the WHI Sequencing Project (phs000281) and the WHI Memory Study (phs000675).

Additional associated genetic resources leveraged include the Population Architecture using Genomics and Epidemiology (PAGE) study (phs000227), the Genomics and Randomized Trials Network (GARNET) study (phs000315), and the SNP Health Association Resource (SHARe) study (phs000386). The primary cohort used for evaluation, the Framingham Heart Study (FHS), is available under accession number phs000007.

## Disclosures of conflicts of interest

The authors declare no competing interests.

## Funding

The research and analysis described in the current publication were supported by a grant (R21MD013681) from the National Institute on Minority Health and Health Disparities and a grant (R01AG080017) from the National Institute on Aging. The funding sponsors were not involved in the study design, data analysis, interpretation of the analysis results, or the manuscript’s preparation, review, or approval.

## Acknowledgment

We sincerely thank the original WHI study investigators and the invaluable participants for their pivotal contributions to advancing women’s health research. We also express our gratitude to the National Institutes of Health (NIH) and the dbGaP for granting access to analyze the WHI data. This work reflects our independent analysis and interpretation and does not represent the views of other parties associated with the WHI study. We sincerely appreciate the collective efforts and contributions of all institutions, collaborators, and teams involved in the WHI study.

The WHI program is funded by the National Heart, Lung, and Blood Institute (NHLBI), NIH, U.S. Department of Health and Human Services through contracts HHSN268201600018C, HHSN268201600001C, HHSN268201600002C, HHSN268201600003C, and HHSN268201600004C. This manuscript was not prepared in collaboration with investigators of the WHI, has not been reviewed or approved by the WHI, and does not necessarily reflect the opinions of the WHI investigators or the NHLBI.

WHISP was funded by Grant Number RC2 HL102924. This study was part of the NHLBI Grand Opportunity Exome Sequencing Project (GO-ESP). Funding for GO-ESP was provided by NHLBI grants RC2 HL103010 (HeartGO), RC2 HL102923 (LungGO) and RC2 HL102924 (WHISP). The exome sequencing was performed through NHLBI grants RC2 HL102925 (BroadGO) and RC2 HL102926 (SeattleGO).

Funding for WHI SHARE genotyping was provided by NHLBI Contract N02- HL-64278.

WHI PAGE is funded through the NHGRI PAGE network (Grant Number U01 HG004790). Assistance with phenotype harmonization, SNP selection, data cleaning, meta-analyses, data management and dissemination, and general study coordination, was provided by the PAGE Coordinating Center (U01HG004801-01).

Funding support for WHI GARNET was provided through the NHGRI GARNET (Grant Number U01 HG005152). Assistance with phenotype harmonization and genotype cleaning, as well as with general study coordination, was provided by the GARNET Coordinating Center (U01 HG005157). Assistance with data cleaning was provided by the National Center for Biotechnology Information. Funding support for genotyping, which was performed at the Broad Institute of MIT and Harvard, was provided by the NIH Genes, Environment and Health Initiative (U01 HG004424).

## Notes

### Competing Interest Statement

The authors have declared no competing interest.

### Author Declarations

Use of WHI and FHS data for the present research was approved by the institutional review board (IRB) at The Ohio State University (protocol STUDY20250826).

