## Supplemental File for "HyTrax: Deep Sequential Modeling of Serial Musculoskeletal Measurements for Fracture Prediction in the Women’s Health Initiative with External Evaluation in the Framingham Heart Study"

### Supplementary Material

**Figure S1.** Study Flow Diagram of WHI Participants Included in the Analysis.

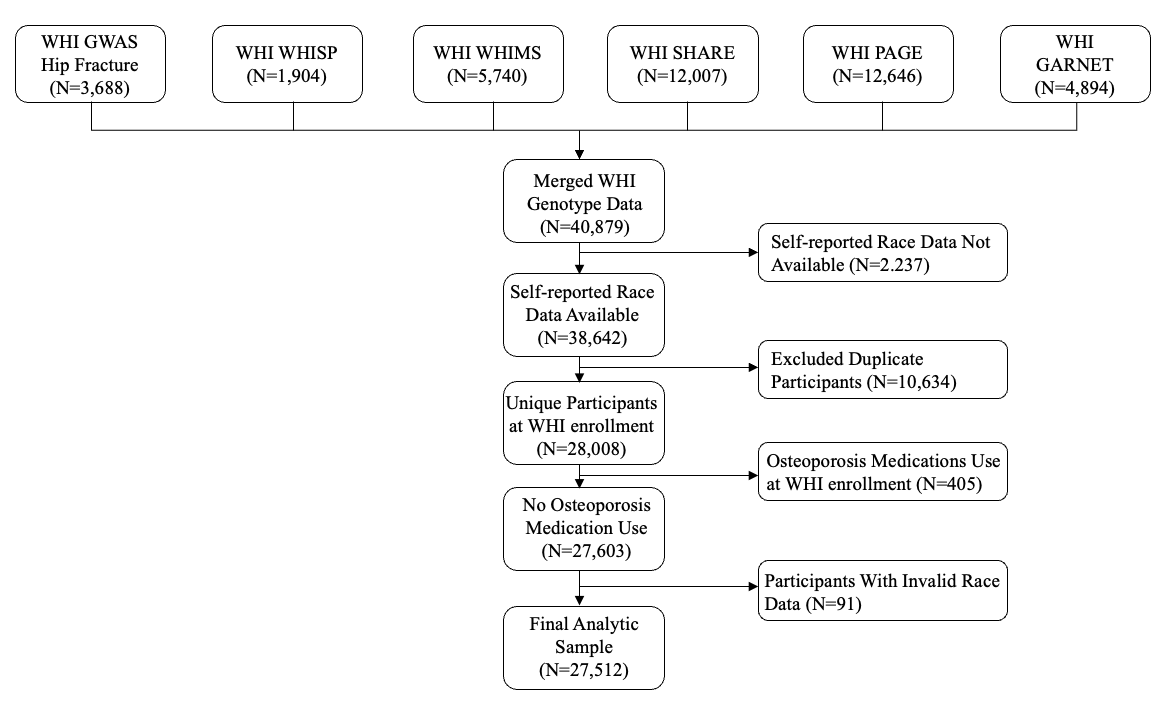

**Abbreviations**: WHI, Women’s Health Initiative; BMD, Bone Mineral Density; GWAS, Genome-Wide Association Study; WHIMS, Women’s Health Initiative Memory Study; WHISP, WHI Sequencing Project; SHARE, SNP Health Association Resource; PAGE, Population Architecture using Genomics and Epidemiology; GARNET, Genomics and Randomized Trials Network.

**Figure S2.** Study Flow Diagram of Framingham Heart Study External Evaluation Cohort Participants Included in the Analysis.

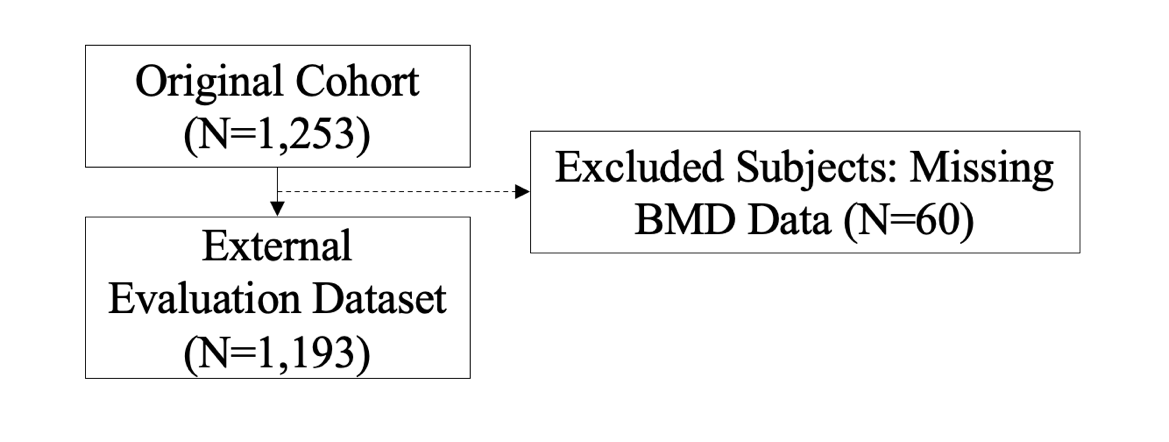

**Figure S3**. **Global Feature Importance of the Hybrid Trajectory-Based (HyTrax) Model Using Shapley Additive exPlanations (SHAP)Values.** (A) Longitudinal features ranked by mean absolute SHAP values. (B) Static (baseline) features ranked by mean absolute SHAP values. The final selected HyTrax model was used to calculate the SHAP value on the held-out WHI internal validation set (N=8,254). For each feature, we calculated the mean absolute SHAP value across all individuals to quantify its overall contribution to prediction.

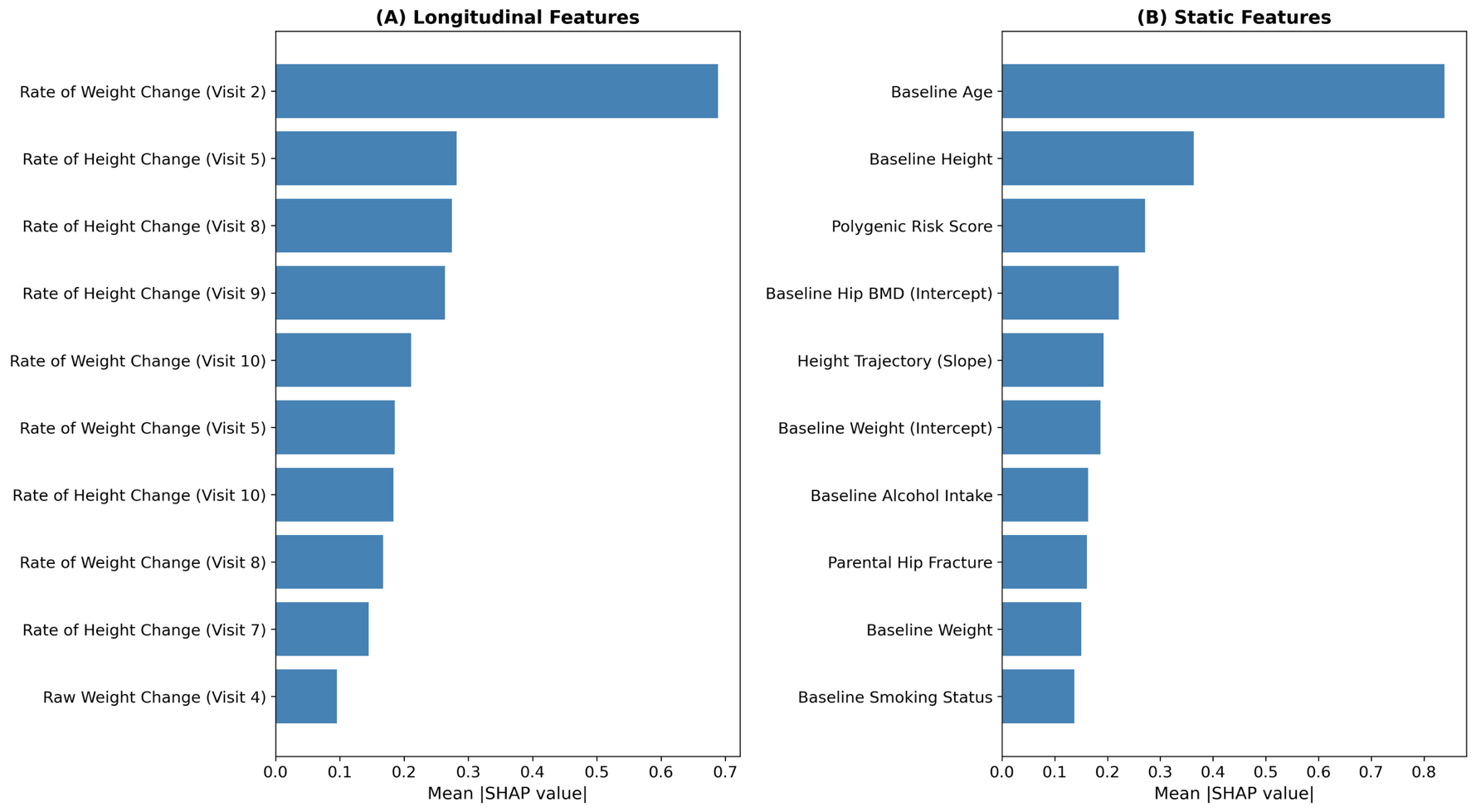

**Notes**: Longitudinal features correspond to tokenized longitudinal measurements provided to the Transformer encoder at successive clinic visits. Static features represent time-invariant demographic characteristics or clinical risk factors assessed at the baseline visit that do not change across the longitudinal sequence.

**Table S1. Overview of Women’s Health Initiative Sub-Studies Included in This Analysis.**

| **Study Name** | **dbGaP* Study Accession Number** | **Characteristic of Study** | **Sample Size** |
| --- | --- | --- | --- |
| Genomics and Randomized Trials Network | phs000315 | - Case-control study within the Genomics and Randomized Trials Network funded by NHGRI - Participants were selected as a nested case-control sample of coronary heart disease, stroke, venous thrombosis, and incident diabetes events. | 4,894 |
| SNP Health Association Resource | phs000386 | - Prospective cohort study aimed at discovering or replicating genes associated with quantitative traits (e.g., blood pressure, lipids) among African American and Hispanic women. - Conducted with WHI participants consenting to Supplemental Consent for broad data sharing. | 12,007 |
| Population Architecture using Genomics and Epidemiology | phs000227 | - Prospective cohort study of genotypes generated via BeadXpress and Metabohip, as part of NHGRI's PAGE project. - Eligibility included postmenopausal women aged 50-79, residing in study areas for at least 3 years post-enrollment. | 12,646 |
| Women’s Health Initiative Memory Study | phs000675 | - Prospective cohort of women aged 65 and older, recruited from the WHI Hormone Trial. - Participants selected for this sub-study include the following WHI Hormone Trial European American women with the appropriate consent for data to be shared on dbGaP. | 5,740 |
| WHI Sequencing Project | phs000281 | - Prospective cohort using next-generation sequencing to identify genes and mechanisms in heart, lung, and blood disorders. - Eligible participants provided DNA samples and consented to data submission to dbGaP. | 1,904 |
| WHI Harmonized and Imputed GWAS Data of Hip Fracture | phs000746 | - Prospective study identifying genetic contributors to hip fracture risk through GWAS among postmenopausal WHI participants. | 3,688 |

*dbGaP: the database of Genotype and Phenotype.

**Table S2. Prediction-Time and Evaluation Design by Cohort.**

| **Design Element** | **WHI (Development & Internal Evaluation)** | **FHS (External Evaluation)** |
| --- | --- | --- |
| Prediction time (t₀) | Individualized: each participant's final recorded clinical visit prior to fracture, death, or censoring | Fixed landmark: Exam 24 (universal across all FHS participants) |
| Allowed input data | All longitudinal measurements strictly before t₀; no post-t₀ data were accessible to the model | Longitudinal measurements from Exams 20, 22, and 24 only; all inputs temporally precede t₀ |
| Event window | 10 years after individualized t₀ | 10 years strictly after Exam 24 |
| Prior fractures | Participants with fractures before t₀ were excluded from the incident fracture analysis | Fractures occurring between Exam 20 and Exam 24 were treated as prevalent events and encoded as a binary "previous fragility fracture" covariate; only fractures strictly after Exam 24 were counted as outcomes |
| Death handling — model fitting | Participants who died before fracture within 10 years were treated as non-events, allowing the algorithm to empirically estimate 10-year cause-specific fracture risk in the presence of competing mortality | For the FHS-local Baseline 1 and 2 comparator models, participants who died before fracture within 10 years were treated as non-events, mirroring the WHI framework. No HyTrax model fitting was performed in FHS. |
| Death handling — evaluation | Model probabilities treated as continuous risk scores; time-dependent metrics with IPCW applied across full follow-up | Model probabilities treated as continuous risk scores; time-dependent metrics with IPCW applied across full follow-up (consistent with WHI). |
| Censoring before 10 years | Participants censored before 10 years were retained in evaluation; IPCW weights dynamically adjusted for right-censoring | Same approach; participants censored before 10 years retained with IPCW adjustment |

**Note:** The individualized t₀ design in WHI emulates a real-world clinical scenario in which a clinician uses all available longitudinal history up to the current visit to generate a 10-year fracture risk estimate. The fixed-landmark design in FHS provides a conservative prospective test of transportability without the advantage of a rolling observation window. In both cohorts, all measurements after t₀ were strictly masked prior to prediction.

**Table S3**. **Frequency Distribution of Longitudinal Measurements for Musculoskeletal Biomarkers in the Women’s Health Initiative (WHI) cohort and FHS Cohorts.** This table displays the number of subjects with a specific count of valid measurements (visits) for each longitudinal variable. For the WHI cohort (N = 27,512), measurements span up to 10 follow-up visits. For the Framingham Heart Study (FHS) evaluation cohort (N = 1,193), measurements were available from up to 3 examination cycles (Exams 20, 22, 24).

| **Dataset** | **Number of Valid Visits** | **Hip BMD (N)** | **Spine BMD (N)** | **Grip Strength (N)** | **Height (N)** | **Weight (N)** |
| --- | --- | --- | --- | --- | --- | --- |
| **WHI** | 0 (None) | 24,843 | 25,107 | 25,434 | 23 | 18 |
|  | 1 | 278 | 264 | 88 | 2,572 | 2,510 |
|  | 2 | 364 | 368 | 199 | 7,857 | 7,906 |
|  | 3 | 716 | 726 | 432 | 1,333 | 1,350 |
|  | 4 | 493 | 507 | 1,306 | 664 | 654 |
|  | 5 | 779 | 764 | 53 | 864 | 858 |
|  | 6 | 39 | 40 | 0 | 1,258 | 1,179 |
|  | 7 | 0 | 0 | 0 | 3,684 | 3,627 |
|  | 8 | 0 | 0 | 0 | 4,740 | 4,795 |
|  | 9 | 0 | 0 | 0 | 2,723 | 2,768 |
|  | 10 | 0 | 0 | 0 | 1,363 | 1,400 |
| **FHS** | 0 (None) | 14 | 175 | 1,176 | 27 | 22 |
|  | 1 | 393 | 367 | 17 | 345 | 322 |
|  | 2 | 331 | 279 | 0 | 316 | 303 |
|  | 3 | 455 | 372 | 0 | 505 | 546 |

**Note:** "0 (None)" indicates participants with no valid measurements for that specific variable across the entire follow-up period. For grip strength in the FHS cohort, the 17 subjects with a single valid measurement were assessed at the later follow-up visit (Exam 24), rather than at the baseline examination (Exam 20); placeholder variables were created for earlier exams to preserve temporal structure.

**Table S4. The List of Hyperparameters Tested and Selected in the** **Hybrid Trajectory-Based (HyTrax) Model**. This table details the hyperparameter search space and the final values selected for the best-performing HyTrax model configuration.

| **Parameter** | **Description** | **Tested Values** | **Final Selected Value** |
| --- | --- | --- | --- |
| **Model Architecture** | | | |
| Embedding Dimension | Dimensionality of the Transformer input embedding and internal layers. | 32, 64 | 32 |
| Number of Attention Heads | Number of parallel attention layers in the Transformer. | 2, 4 | 2 |
| Number of Layers | Depth of the Transformer Encoder. | 1, 2 | 1 |
| Dropout Rate | Dropout applied in the Transformer encoder and prediction head (for regularization). | 0, 0.1 | 0 |
| **Training Parameters** | | | |
| Final Epochs | Total epochs used for final training of the selected model. | 200 | 200 |
| Learning Rate | Step size for the Adam optimizer. | $\text{5 × }\text{10}^{\text{-4}}$ | $\text{5 × }\text{10}^{\text{-4}}$ |
| Batch Size | Number of samples processed per gradient update. | 128 | 128 |
| Gradient Clipping | Maximum value for gradient norm to prevent exploding gradients. | 1.0 | 1.0 |
| **Cross-Validation** | | | |
| Cross-Validation Folds | Number of folds used for the initial hyperparameter search. | 5 | 5 |

**Table S5. Definitions of Predictive Models and Ensembles Evaluated in the Framingham Heart Study (FHS) External Evaluation Cohort.**

| **Model Name** | **Model Type** | **Included Static/Baseline Predictors** | **Included Longitudinal Predictors** |
| --- | --- | --- | --- |
| Baseline 1 (FRAX-CRF Surrogate) | Logistic Regression | Age, sex, height, weight, smoking status, alcohol intake, previous fragility fracture, rheumatoid arthritis, and glucocorticoid use. | None |
| Baseline 2 (Extended Clinical + BMD) | Logistic Regression | All features from Baseline 1 plus: Total hip BMD, lumbar spine BMD, Polygenic Risk Score (PRS), and grip strength. | None |
| HyTrax | Transformer | Subject-specific mixed-effects slopes and static clinical covariates. | Tokenized longitudinal trajectories of: Hip/spine BMD, grip strength, weight, and height. |
| HyTrax + Baseline 1 | Weighted Ensemble | All features from Baseline 1. | All trajectories from HyTrax. |
| HyTrax + Baseline 2 | Weighted Ensemble | All features from Baseline 2. | All trajectories from HyTrax. |

**Abbreviations**: BMD, Bone Mineral Density; CRF, Clinical Risk Factor; FHS, Framingham Heart Study; FRAX, Fracture Risk Assessment Tool; PRS, Polygenic Risk Score.

**Notes**: * Parental hip fracture history: While a standard component of the FRAX algorithm, this variable was omitted from the baseline models in this external evaluation due to data unavailability in the FHS dbGaP dataset.

- Ensemble Models: The "HyTrax +" models represent weighted ensembles. They integrate the individualized, dynamic risk embedding generated by the longitudinal Transformer (HyTrax) with the static probabilities generated by the respective baseline logistic regression models to maximize predictive performance.

**Table S6**. **Comparison of Baseline Demographic and Clinical Characteristics of Women’s Health Initiative Participants, Stratified by Hip Fractures Event (N=27,512).**

| **Variable** | **Participants without Hip Fractures (N=25,715)** | **Participants with Hip Fractures (N=1,797)** | **P-value*** |
| --- | --- | --- | --- |
| Age (years), mean (SD) | 63.3 (7.4) | 69.5 (6.4) | <0.001 |
| Height (cm), mean (SD) | 161 (6.3) | 162 (6.5) | <0.001 |
| Weight (kg), mean (SD) | 77.2 (17.1) | 69.6 (14.5) | <0.001 |
| FRAX for Hip, mean (SD) | 1.9 (3.8) | 5.6 (6.5) | <0.001 |
| FRAX (with BMD) for Hip, mean (SD) | 1.4 (2.9) | 4.5 (5.1) | <0.001 |
| Hip BMD (g/cm^2^), mean (SD) | 0.9 (0.2) | 0.8 (0.1) | <0.001 |
| Spine BMD (g/cm^2^), mean (SD) | 1.0 (0.1) | 0.9 (0.1) | <0.001 |
| Grip Strength (kg), mean (SD) | 23.2 (6.2) | 22.7 (6.2) | 0.07 |
| PRS ^a^, mean (SD) | 2.2 (22.9) | 7.0 (23.2) | <0.001 |
| RACE/ethnicity, $n (\%)$ |  |  | <0.001 |
| American Indian or Alaskan Native | 582 (2.3%) | 9 (0.5%) |  |
| Asian or Pacific Islander | 484 (1.9%) | 15 (0.8%) |  |
| African American | 9,999 (38.9%) | 56 (3.1%) |  |
| Hispanic/Latino | 4,607 (17.9%) | 27 (1.5%) |  |
| Non-Hispanic White | 10,043 (39.1%) | 1,690 (94.0%) |  |
| Current Smoking, $n (\%)$ | 2,433 (9.5%) | 146 (8.1%) | 0.03 |
| Rheumatoid Arthritis, $n (\%)$ | 3,419 (13.3%) | 252 (14.0%) | 0.41 |
| Previous fragility fracture, $n (\%)$ | 314 (1.2%) | 74 (4.1%) | <0.001 |
| Parental hip fracture history, $n (\%)$ | 812 (3.2%) | 110 (6.1%) | 0.05 |
| Alcohol intake, $n (\%)$ | 16,722 (65.0%) | 1,285 (71.5%) | <0.001 |
| Glucocorticoid use, $n (\%)$ | 39 (0.2%) | 9 (0.5%) | 0.002 |
| Diabetes, $n (\%)$ | 2,282 (8.9%) | 118 (6.6%) | 0.44 |
| ≥ 1 Previous falls, $n (\%)$ | 8,250 (32.1%) | 714 (39.7%) | <0.001 |

**Abbreviations**: FRAX, Fracture Risk Assessment Tool; SD, Standard Deviation; BMD, Bone Mineral Density; PRS, Polygenic Risk Score.

^*^ P-value was obtained by t-test for continuous variables and chi-square tests for the categorical variable.

^a^ Developed using the LDpred algorithm with 103,155 genetic variants from UK Biobank GWAS summary statistics.

**Table S7**. **Comparison of Baseline Demographic and Clinical Characteristics Between the Development Cohort (WHI, N=27,512) and the External Evaluation Cohort (FHS, N=1,193).** The FHS evaluation subset consisted exclusively of Non-Hispanic White participants, precluding statistical comparison for certain racial/ethnic subgroups.

| **Variable** | **WHI**  **(N=27,512)** | **FHS**  **(N=1,193)** | **P-value*** |
| --- | --- | --- | --- |
| Age (years), mean (SD) | 63.7 (7.5) | 75.3 (4.9) | <0.001 |
| Height (cm), mean (SD) | 161 (6.4) | 157 (9.6)^a^ | <0.001 |
| Weight (kg), mean (SD) | 76.7 (17.1) | 69.9 (13.9)^a^ | <0.001 |
| Hip BMD (g/cm^2^), mean (SD) | 0.9 (0.2) | 0.7 (0.1) | <0.001 |
| Spine BMD (g/cm^2^), mean (SD) | 1.0 (0.1) | 0.9 (0.1) | <0.001 |
| Grip Strength (kg), mean (SD) | 23.1 (6.2) | NA^b^ | NA |
| PRS^c^, mean (SD) | 4.3 (25.4) | 6.4 (18.1) | <0.001 |
| RACE/ethnicity, $n (\%)$ |  |  | <0.001 |
| American Indian or Alaskan Native | 591 (2.1%) | NA |  |
| Asian or Pacific Islander | 499 (1.8%) | NA |  |
| African-American | 10,055 (36.5%) | NA |  |
| Hispanic/Latino | 4,634 (16.8%) | NA |  |
| Non-Hispanic White | 11,733 (42.6%) | 1,193 (100.0%) |  |
| Current Smoking, $n (\%)$^e^ | 2,579 (9.4%) | 13 (1.7%) | <0.001 |
| Rheumatoid Arthritis, $n (\%)$^e^ | 3,671 (13.3%) | 57 (7.5%) | <0.001 |
| Alcohol intake, $n (\%)$^e^ | 18,007 (65.5%) | 492 (57.7%) | <0.001 |
| Glucocorticoid use, $n (\%)$^e^ | 48 (<1%) | 24 (3.1%) | <0.001 |
| ≥ 1 Previous fall, $n (\%)$^e^ | 8,964 (32.6%) | 179 (23.5%) | <0.001 |
| Major Osteoporotic Fracture^d^, $n (\%)$ | 2,597 (9.4%) | 173 (14.5%) | <0.001 |
| Hip Fractures, $n (\%)$ | 1,797 (6.5%) | 120 (10.1%) | <0.001 |

**Abbreviations**: SD, Standard Deviation; BMD, Bone Mineral Density; PRS, Polygenic Risk Score.

^*^ P-value was obtained by t-test for continuous variables and chi-square tests for the categorical variable.

^a^ Height and weight in the FHS cohort were originally recorded in inches and pounds; these were converted to centimeters and kilograms, respectively, to harmonize scales with the WHI dataset.

^b^ Grip strength is reported as not applicable (NA) for the FHS cohort in this table because measurements were not collected at the baseline examination (Exam 20). In the FHS dataset, grip strength was only assessed at a later follow-up visit (Exam 24) for a very small subset of participants (n=17); placeholder variables were created for earlier exams to preserve temporal structure.

^c^ Developed using the LDpred algorithm with 103,155 genetic variants from UK Biobank GWAS summary statistics.

^d^ Major Osteoporotic Fracture was defined as a fracture occurring at the hip, spine, shoulder, or wrist.

^e^ FHS percentages for these variables are calculated among participants with non-missing data, which may be fewer than the total N=1,193 due to item-level missingness. Denominators were as follows: Current Smoking n=752; Rheumatoid Arthritis n=760; Alcohol intake n=853; Glucocorticoid use n=774; Previous fall n=762. Percentages calculated using N=1,193 as the denominator would differ from those shown.
